# Estimating and visualising multivariable Mendelian randomization analyses within a radial framework

**DOI:** 10.1101/2023.04.04.23288134

**Authors:** Wes Spiller, Jack Bowden, Eleanor Sanderson

## Abstract

**Background:** Multivariable Mendelian randomization (MVMR) is a statistical approach using genetic variants as instrumental variables to estimate direct causal effects of multiple exposures on an outcome simultaneously. In univariable MR findings are typically illustrated through plots created using summary data from genome-wide association studies (GWAS), yet analogous plots for MVMR have so far been unavailable due to the multidimensional nature of the analysis.

**Methods:** We propose a radial formulation of MVMR, and an adapted Galbraith radial plot, which allows for the direct effect of each exposure within an MVMR analysis to be visualised. Radial MVMR plots facilitate the detection of outlier variants, indicating violations of one or more assumptions of MVMR. In addition, the RMVMR R package is presented as accompanying software for implementing the methods described.

**Results:** We demonstrate the effectiveness of the radial MVMR approach through simulations and applied analyses, estimating the effect of lipid fractions on coronary heart disease (CHD). We find evidence of a protective effect of high-density lipoprotein (HDL) and a positive effect of low-density lipoprotein (LDL) on CHD, however, the protective effect of HDL appeared to be smaller in magnitude when removing outlying variants. In combination with simulated examples, we highlight how important features of MVMR analyses can be explored using a range of tools incorporated within the RMVMR R package.

**Conclusions:** Radial MVMR effectively visualises causal effect estimates, and provides valuable diagnostic information with respect to the underlying assumptions of MVMR.

## Introduction

Mendelian randomization (MR) is a methodological framework in which genetic variants-often single nucleotide polymorphisms (SNPs)- are used as instrumental variables to estimate causal relationships in the presence of unmeasured confounding [1]. MR analyses are often performed using summary data from publicly available genome-wide association studies (GWAS), reflecting the ease with which such data can be accessed in contrast with individual-level data [2, 3].

A typical summary MR study begins with identifying a set of SNPs associated with the exposure of interest, after which SNP-exposure and SNP-outcome association estimates for each SNP are obtained [2, 3]. Individually, dividing the SNP-outcome association by the SNP-exposure association yields a Wald ratio estimate for the effect of the exposure on the outcome corresponding to each SNP [2, 4]. When multiple SNPs are used ratio estimates are typically combined using inverse variance weighting (IVW), producing an average causal effect estimate. SNPs are often weighted by the inverse of the variance of their SNP-outcome association, though a range of weighting specifications can be applied [2, 5, 6].

The extent to which MR causal effect estimates are unbiased is largely determined by three key assumptions. SNPs serving as instruments must be robustly associated with the exposure of interest (IV1), independent of confounders of the exposure and outcome (IV2), and independent of the outcome when conditioning on the exposure (IV3) [7]. Assumption IV1 requires the denominator (the SNP-exposure association) to be non-zero, ensuring the ratio estimate is defined. Assumptions IV2-3 require the SNP-outcome association to be the product of the SNP-exposure association and causal effect of interest, such that the association between the SNP and outcome is entirely mediated by the exposure of interest (see supplementary material). As ratio estimates using valid (IV1-3 satisfying) SNPs would asymptotically converge towards the causal effect of interest, observed heterogeneity in effect estimates using many SNPs can potentially serve as an indicator of IV2-3 violation.

The value of estimated heterogeneity as an indicator of IV2-3 violation serves as the motivation for conducting summary MR analyses within a radial framework, previously described in Bowden et al (2018) [6]. Radial MR adapts the original summary MR regression model such that causal estimates are a function of the ratio estimate and weighting corresponding to each SNP. Radial IVW, for example, regresses the product of the ratio estimate and square root weighting for each SNP upon the set of square root weightings, omitting an intercept. This produces an IVW causal effect estimate identical to the standard IVW approach, while allowing effects to be visualised using an adapted Galbraith radial plot [10, 11]. Importantly, such plots show the weighting attributed to each SNP on the x-axis, while the contribution to global heterogeneity is proportional to the distance of each data point from the superimposed regression line, or its estimated residual. This facilitates outlier detection, highlighting SNPs which may violate the underlying MR assumptions. Finally, as the weighting applied to each SNP is always positive, and the ratio estimates themselves are independent of allele coding, there is no need to reorient SNP-exposure associations [6].

SNP-outcome associations which violate assumptions IV2-3 can potentially be mediated through additional phenotypes for which measures are available, and in such cases multivariable Mendelian randomization (MVMR) approaches can be used to estimate the direct effect of multiple exposures on an outcome simultaneously through a generalisation of the univariable IVW model [3, 12–14]. Such analyses are particularly valuable where SNPs selected as instruments in univariable analyses violate assumptions IV2 or IV3 through a measured phenotypic pathway, as such associations can be accounted for when estimating causal effects. In the summary data setting, Burgess et al (2015) demonstrate how MVMR estimates can be obtained using a generalisation of the IVW model, specifically by regressing SNP-outcome associations upon SNP-exposure associations obtained for each included exposure [3, 12]. Sanderson et al (2019) further develop these methods, proposing a range of sensitivity analyses specific to MVMR. This includes methods to assess conditional instrument strength (an extension of IV1 necessary for MVMR analyses) and horizontal pleiotropy (IV3) [13, 14]. However, while several pleiotropy robust methods for MVMR have been proposed, there is an absence of approaches to effectively visualise MVMR analyses [15, 16].

In this paper we present a radial MVMR approach which allows for important features of conventional MVMR analyses to be highlighted, in particular SNPs which violate the underlying assumptions of MVMR. Radial MVMR addresses two key limitations of existing approaches, providing a means with which MVMR analyses can be visualised and a process through which outliers can be detected after conditioning on additional exposures. Initially, we demonstrate how univariable radial MR can be extended to incorporate multiple exposures, creating a radial analogue of the IVW MVMR model. With this complete we describe how MVMR estimates can be visualised using radial plots, and crucially, how including an adjustment to ratio estimates to account for additional exposures included in the MVMR model facilitates the detection of pleiotropic SNPs. Specifically, outliers in a radial MVMR analysis can be formally and visually identified through an evaluation of their contribution to global heterogeneity, indicating likely violations of assumption IV3. Through simulated analyses we also highlight the extent to which pruning for such outliers can greatly improve causal effect estimation, both in terms of reducing observed bias and increasing the precision of MVMR estimates.

To demonstrate the application of radial MVMR we present an applied example evaluating the effects of low-density lipoprotein (LDL), high-density lipoprotein (HDL), and triglycerides on coronary heart disease (CHD). Using publicly available summary data from the Global Lipids Genetics Consortium (GLGC) and CARDIoGRAMplusC4D Consortium we find evidence of a protective effect of HDL, and a positive causal effect of LDL in relation to CHD [17, 18]. When pruning for identified outliers the effect of HDL decreases in magnitude, and we find evidence of a positive association with both LDL and triglycerides. We also illustrate how pleiotropic bias appears likely when conducting univariable analyses, and how such bias is potentially mitigated when including additional exposures. Throughout we perform all analyses using the RMVMR R package for the R software environment, which has been developed to facilitate the application of radial MVMR analyses. The RMVMR R package is freely available from https://github.com/WSpiller/RMVMR.

## Methods

### Univariable summary Mendelian randomization

In univariable MR analyses one or more SNPs are used as instruments to estimate the causal effect of a single exposure *X* upon an outcome *Y*. Let *G*_*j*_ represent the *j*^*th*^ independent SNP from a set of *j* ∈ {1, 2, …, *J*}, and let *U* denote one or more unmeasured confounders. A SNP is considered valid provided it satisfies assumptions IV1-3, with assumed relationships depicted in Fig 1. Letting *i* ∈ {1, 2, …, *N*} index subjects:

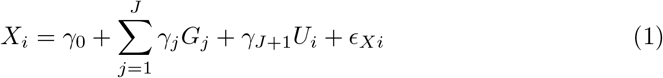

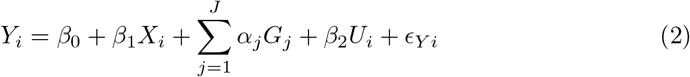

If all confounders of the exposure and outcome were measured, an unbiased estimate for the effect of *X* upon *Y* (*β*_1_) could be estimated by performing a multivariable regression of *Y* upon *X* including all confounders in the set *U*. As information on *U* is unavailable by definition, summary MR uses SNP-exposure and SNP-outcome associations obtained for each SNP to estimate the effect of *X* on *Y* in a manner robust to confounding bias. SNP-exposure and SNP-outcome associations are estimated using the following simple regression models (excluding variables commonly included in GWAS such as principal components or age).

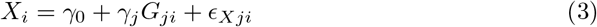

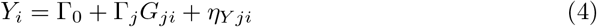

Using the estimated total effect of *G*_*j*_ on 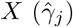 and the estimated total effect of *G*_*j*_ on *Y* (Γ_*j*_), the ratio estimate for a given SNP 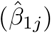 using estimated parameters 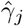 and 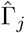 can be written as:

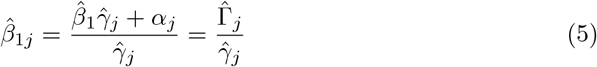

Assumptions IV1-3 imply *γ*_*j*_ ≠ 0 and *α*_*j*_ = 0, and consequently the ratio can be shown to be a consistent estimator of the effect *β*_1_ provided assumptions IV1-3 hold (see supplementary material). Using *w*_*j*_ to represent the weight applied to each SNP *j*, the IVW estimate using multiple uncorrelated SNPs is given by

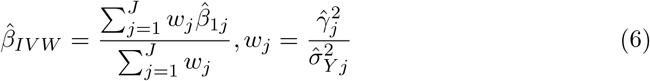

Note that in Eq 6 inverse variance weights are used, though a wide-range of weightings are available. As described in Bowden et al (2018), an equivalent IVW estimate in Eq 6 can be obtained by fitting a radial regression model, regressing the product of the ratio estimate and square root weight attributed to each SNP against the set of square root weights across all SNPs [6].

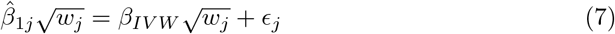

Constructing a scatter plot with 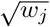 and 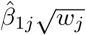 on the y-axis, and superimposing the regression line from Eq 7, the distance of each observation from the regression line is equal to its square-root contribution to Cochran’s heterogeneity statistic, 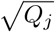, where

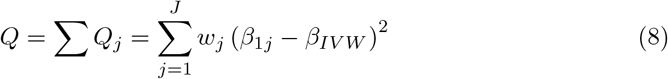

As previously described in Bowden et al (2018), the global Q-statistic follows a chi-squared distribution with *J* − 1 degrees of freedom, and individual estimates *Q*_*j*_ have a chi-squared distribution with 1 degree of freedom [6]. This allows p-values to be used as a threshold for identifying outlying SNPs. Fig 2 shows an example radial plot constructed using previously published GWAS summary data from Do et al, included within the RadialMR R package [17]. This data contains information on LDL from the Global Lipids Genetics Consortium (GLGC), and CHD data from the CARDIoGRAM study [19, 20]. Considering the effect of LDL upon CHD, variants identified as outliers are highlighted in yellow and the IVW estimate is represented by a black regression line through the origin.

**Fig 1.**
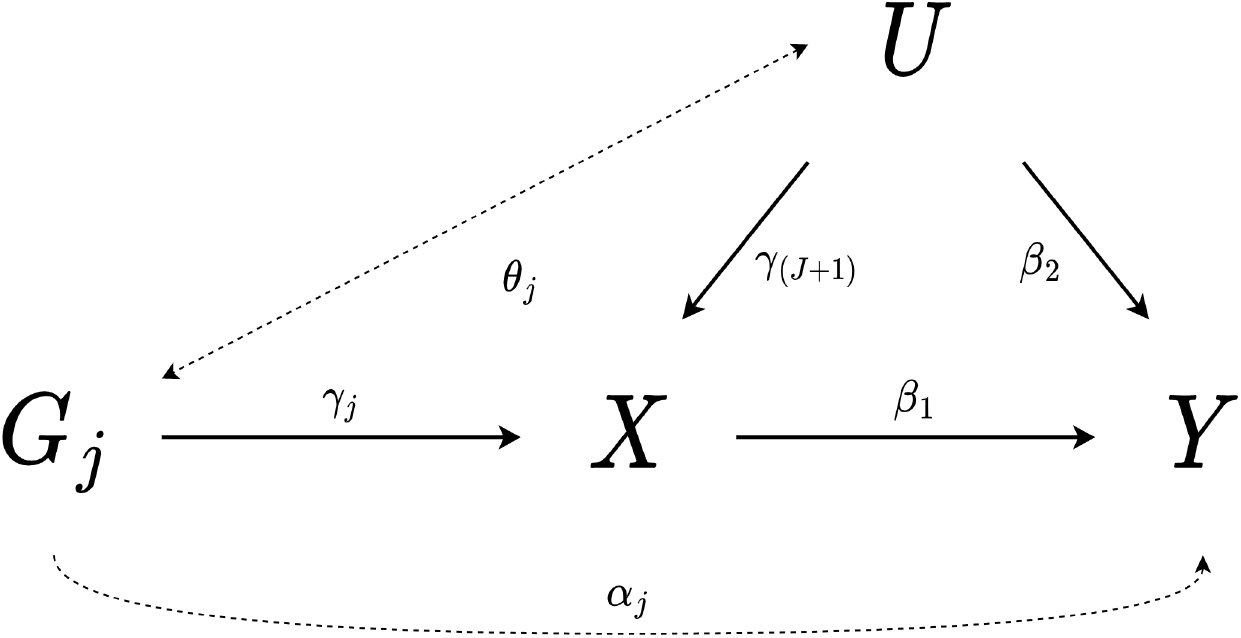
Directed acyclic graph (DAG) illustrating the assumptions of MR. Associations required to be zero for the MR assumptions to be satisfied are shown as dashed lines. Specifically, IV2 is satisfied when *θ*_*j*_ is zero, and IV3 is satisfied when *α*_*j*_ is zero.

**Fig 2.**
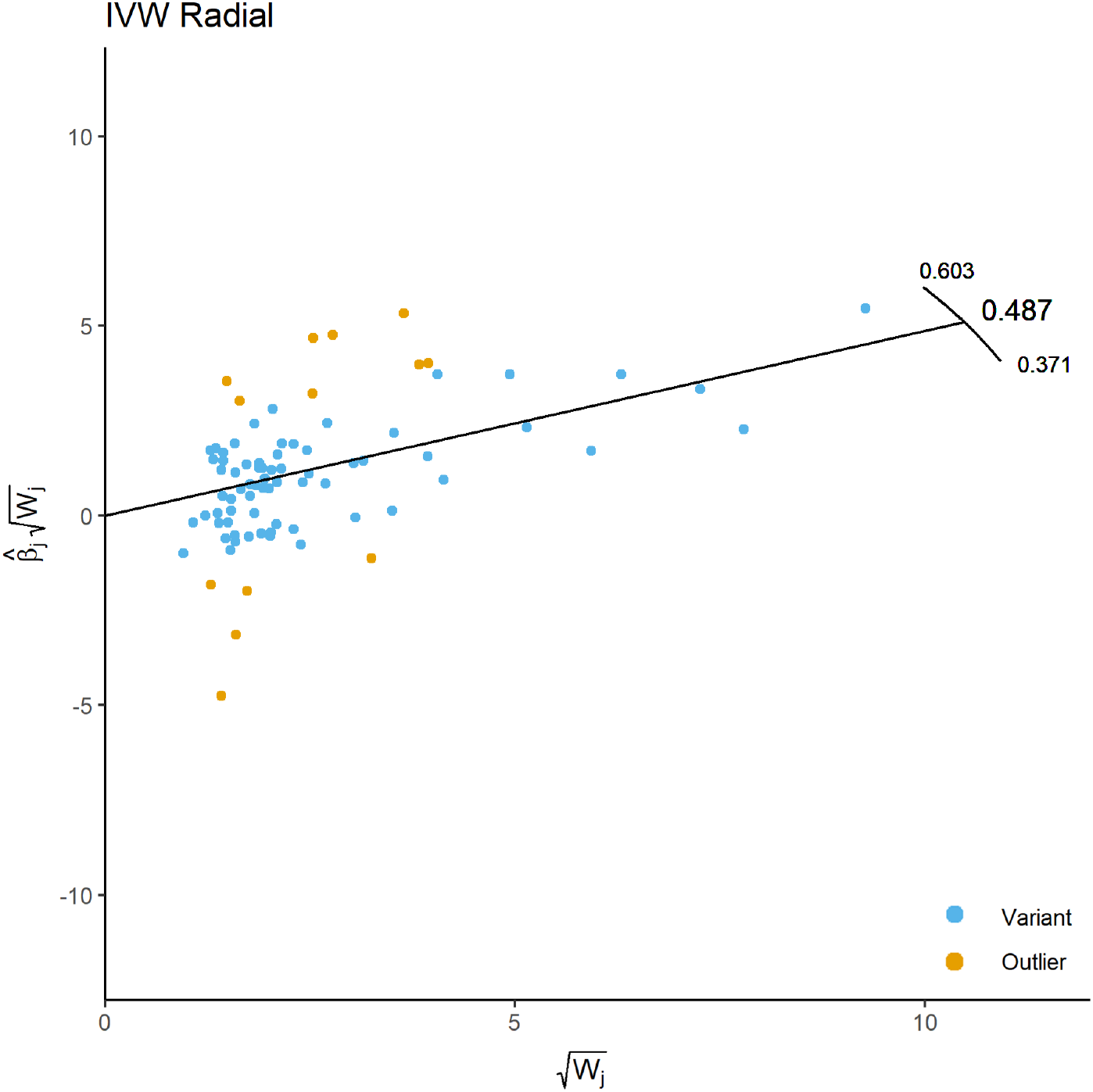
A scatter plot showing a radial IVW estimate using data from Do et al. Here the x-axis represents the square root of the weight applied to each SNP, while the y-axis shows the product of the ratio estimate and square root of the weight given to each SNP. Outliers are identified using Cochran’s Q-statistic and a p-value threshold of 0.05/number of SNPs has been used to correct for multiple testing.

### Multivariable Mendelian randomization

MVMR extends the univariable MR framework to include multiple potentially correlated exposures, leveraging the entire set of SNPs associated with at least one included exposure [13, 14]. This allows for the direct effect of each exposure to be consistently estimated, (that is, the effect of an exposure holding the others fixed), provided the total set of SNPs *G* is:

- Strongly associated with each exposure when conditioning on remaining exposures (MVMR1)
- Independent of all confounders of any individual exposure and the outcome (MVMR2)
- Independent of the outcome when conditioning on all included exposures and all confounders (MVMR3)

The previous data generating model can readily be generalised to include an arbitrary number of SNPs and exposures, though there needs to be at least as many SNPs as exposures for the MVMR model to be identified [13]. For *i* ∈ {1, 2, …, *N*} observations including *j* ∈ {1, 2, …, *J*} SNPs, a set of SNPs *l* ∈ {1, 2, … *L*} where *j* ∉ *l*, a set of exposures *k* ∈ {1, 2, …, *K*}, and a set of exposures *m* ∈ {1, 2, …, *M*} for which *k* ∉ *m*:

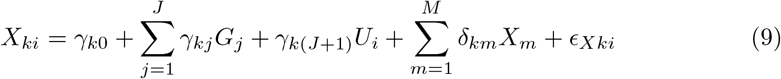

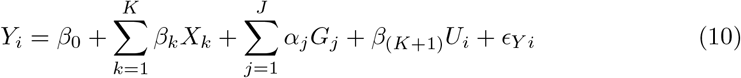

The set of relationships between variables is illustrated in Fig 3.

**Fig 3.**
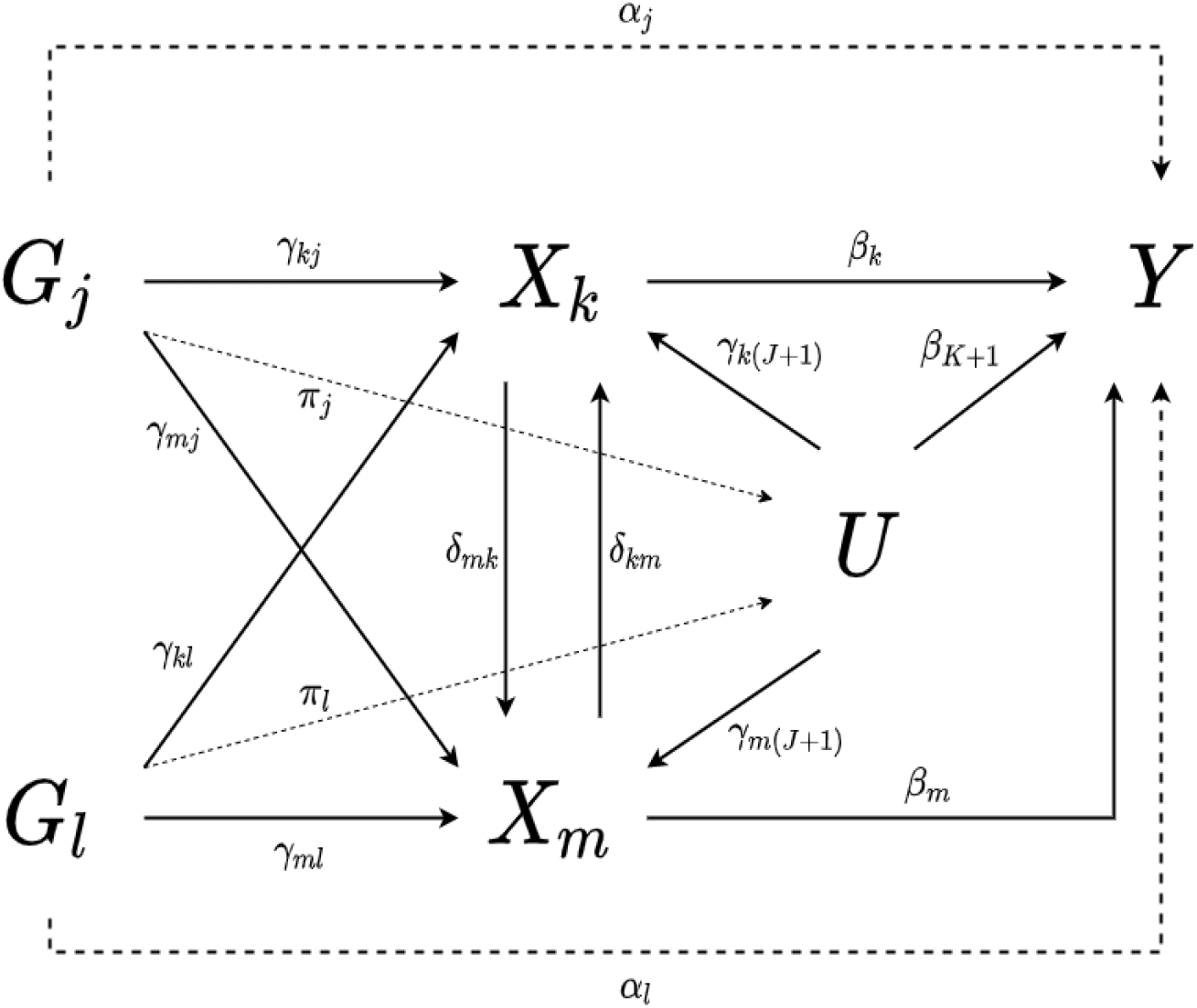
A DAG illustrating associations described in equations 1-2 and 9-10 for an arbitrary number of SNPs and exposures. Dashed lines represent associations which would violate assumptions MVMR2-3.

Using equations (9) and (10) the univariable estimand can be derived. For clarity, we denote the total effect of an instrument *G*_*j*_ on an exposure *X*_*k*_ as 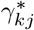, while *γ*_*kj*_ represents the direct effect of *G*_*j*_ on *X*_*k*_ conditioning on all relevant exposures on the pathway from *G*_*j*_ to *X*_*k*_. This allows us to define the univariable ratio estimand as:

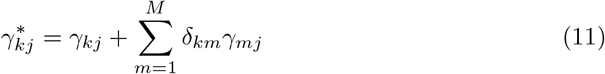

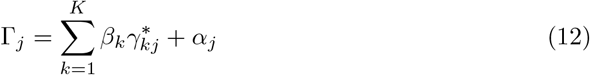

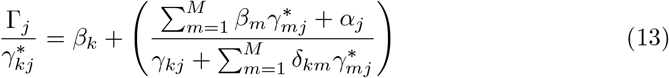

A derivation of this result is provided in the supplementary material. If the MVMR assumptions are satisfied, and independent SNPs are used, Eq 13 simplifies to

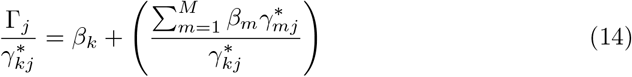

Note that the term in parentheses in Eq 14 represents the effect of the additional exposures *X*_*m*_, and including ratio estimates for all additional exposures *X*_*m*_ within a multivariable regression will yield marginal effects of each exposure, adjusting for this term in each case. In univariable MR, pleiotropic effects violating IV3 would likely be present when 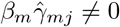. The direct effect of each exposure can be estimated by regressing instrument-outcome associations upon instrument-exposure associations for each exposure simultaneously [2], such that:

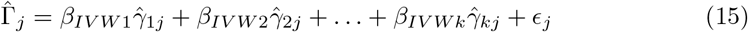

MVMR relies upon a sufficient proportion of instruments being strongly associated with each exposure, conditional on remaining included exposures. This can be evaluated by calculating the conditional F-statistic for each exposure using a conventional threshold of 10 [14]. In terms of ratio estimates the conditional instrument strength of instrument *G*_*j*_ can be thought of as the extent to which 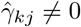, and the conditional independence of the genetic instruments with respect to the outcome (MVMR3) can be evaluated by estimating observed heterogeneity across the set of ratio estimates, as described in Sanderson et al (2021) [14].

### Radial MVMR

The univariable radial MR model can be readily extended to include multiple exposures, creating an analogue of the MVMR regression model shown in Eq 16:

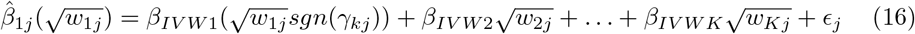

where *w*_*kj*_ represents the weighting for each SNP with respect to exposure *k*. For example, *w*_1*j*_ is equal to 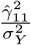 when first order weights are used. It is important to note that the SNP-exposure associations must be re-oriented so as to match the direction of the SNP-exposure associations contributing to the regressand, in contrast to univariable radial MVMR which does not require SNP-exposure associations to be reoriented. This does not alter effect estimates obtained, and they remain equal to those obtained using conventional MVMR.

An immediate benefit of conducting MVMR within a radial framework is plotting 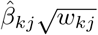 against 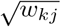 can be accomplished using generalised axes scales, allowing ratio estimates for each exposure to be projected onto the same scatter plot simultaneously. The RMVMR plot has the same x-axis scale as the univariable Radial MR plot, plotting 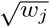 values. The y-axis of the RMVMR plot, however, represents the product of the ratio estimate and weighting for the reference exposure. As all instruments associated with at least one exposure are used to estimate causal effects, an RMVMR plot will have *K* × *J* observations, including a set of weightings for each included exposure *k*.

For clarity, while all weightings are used to estimate causal effects it is often appropriate to limit the number of observations represented on an RMVMR plot to instruments exceeding a given weighting threshold. For example, points corresponding to an exposure *X*_1_ would be shown on the RMVMR plot provided they have an F-statistic greater than 10. This omits clusters of instruments with negligible weightings which are unlikely to be of interest, while improving the readability of the plot.

RMVMR plots are particularly useful as a tool for highlighting the extent to which individual SNPs contribute towards global heterogeneity with respect to each included exposure. Following the the data generating model given in equations Eq 9-10 performing a univariable MR analyses will result in biased estimates where SNPs are associated with multiple causally relevant phenotypes. This is primarily because the sum of associations 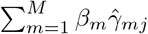 represent pleiotropic pathways through the omitted exposures, resulting in increased heterogeneity provided such effects are not identically distributed across the set of SNPs (see Eq 14). If this is the case, then it follows that adjusting for such associations would result in a decrease in effect estimate heterogeneity, with estimates converging towards the MVMR estimate once the univariable pleiotropic bias is corrected. The radial analogue for Eq 14 can be written as:

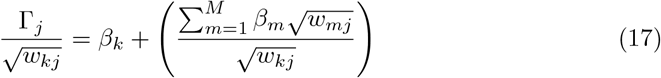

This result has important implications in terms of visualising heterogeneity in RMVMR analyses. In an RMVMR plot we plot the product of the ratio estimate and corresponding square root weighting against each set of weights on a generalised x-axis 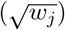. However, as the univariable ratio estimate for each instrument is used, superimposed regression lines representing the RMVMR estimate for each exposure will not represent the best fit through the plotted observations. This is because the ratio estimates do not account for the adjustment from other exposures. We can write the position of each data point on the y-axis as:

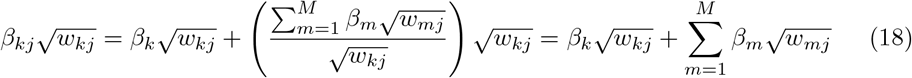

Eq 18 highlights how, by subtracting 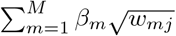 from the y-axis value of each data point, an adjustment can be performed to account for exposures included in the RMVMR model. Crucially, though the true value of *β*_*m*_ is unknown in Eq 18, an adjustment can be made using the estimate of *β*_*m*_ obtained from the RMVMR model, that is, *β*_*IV W m*_ in Eq 16.

When the MVMR assumptions are satisfied MVMR ratio estimates will be unbiased. This means that adjusted ratio estimates should converge towards their corresponding estimated effect, for example, *β*_*k*_ in equation (17). Consequently, observed heterogeneity in MVMR ratio estimates can be indicative of violations of MVMR3. This can be formally evaluated through an adapted form of Cochran’s Q-statistic calculated for each exposure:

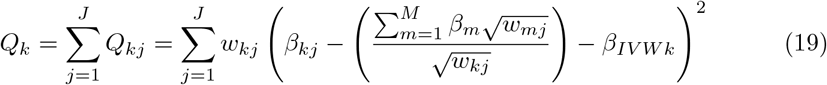

As in the univariable Radial MR setting, the square root contribution of each instrument to global heterogeneity with respect to an exposure *k* is equal to:

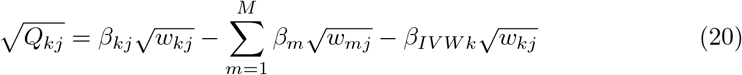

Equation (20) describes how the square root contribution to heterogeneity with respect to exposure *k* is represented by the distance from each adjusted point to the superimposed regression line for *β*_*IVWk*_, evaluated at 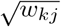.

To visualise the extent to which the addition of an exposure minimises effect estimate heterogeneity, a pair of RMVMR plots can be constructed. Initially an RMVMR plot is created including regression lines showing the MVMR estimate for each exposure. For the first plot, each data point shows the the square root weighting for each instrument 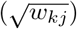, and the product of the square root weighting and unadjusted ratio estimate for each exposure. The second RMVMR plot includes the adjustment to each univariable ratio estimate, described in equation (18). An example of such plots using simulated data is presented in Figure 4.

**Fig 4.**
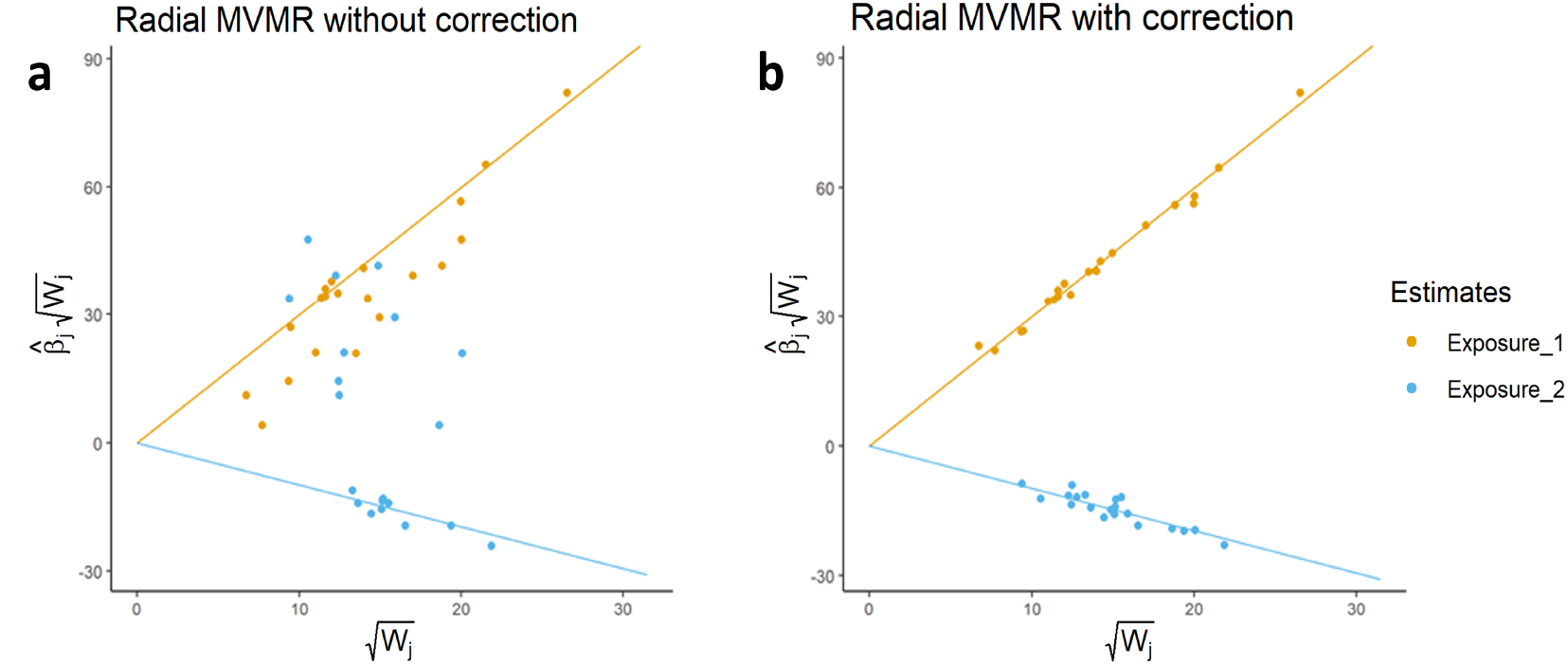
A pair of RMVMR plots using two exposures presented for illustration. In this case, both exposures have a non-zero effect on the outcome, and 10 instruments are associated with both exposures simultaneously. Fig 4a uses ratio estimates prior to adjusting for the additional exposure, resulting in substantial observed heterogeneity. Fig 4b shows a substantial reduction in observed heterogeneity when adjusting for the additional exposure.

In this simulated example a total of 30 instruments are used, of which 10 are associated with exposure *X*_1_, 10 are associated with exposure *X*_2_, and a final 10 instruments are associated with *X*_1_ and *X*_2_ simultaneously. In univariable analyses, provided both exposures have an effect on the given outcome 10 instruments will violate IV3 through the omitted exposure. This is shown by the degree of observed heterogeneity in Figure 4a prior to using adjusted ratio estimates. As shown in Figure 4b, where the inclusion of an additional exposure accounts for remaining pleiotropic effects, the resulting adjusted ratio estimates will converge to the MVMR estimate, that is, the direct effect of each exposure on the outcome of interest.

A number of important features of can be discerned from Figure 4 which warrant consideration. Initially, the degree to which the position of data points changes can serve to indicate whether the inclusion of an additional exposure within an RMVMR model is appropriate. Specifically, if omitting an exposure induces bias when calculating ratio estimates, we would expect the vertical position of data points to change when applying an adjustment. In cases where data points do not appreciably change in position, this can imply either that the additional exposure has no effect on the outcome (*β*_*m*_ = 0), that instruments are exposure specific such that *w*_*mj*_ = 0, or an unlikely scenario where the adjustments across all exposures are balanced such that 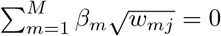.

A second benefit of RMVMR plots is the ability to visually identify the relative exposure-specific weighting of each instrument. In Figure 4 the position of each instrument on the x-axis reflects its weighting 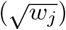 with respect to each individual exposure. It should be noted, however, that this does not reflect the conditional instrument strength of each instrument.

Finally, as highlighted in equation (20) the vertical distance of each data point in Figure 4b from their corresponding superimposed regression line is equal to their square root contribution to heterogeneity with respect to the given exposure. This can be indicative of invalid instruments, and would warrant further follow-up using external data. It is, however, critical to note that the adjustments made to each exposure are reliant upon initial estimates for the direct effect of each exposure 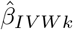. In cases where these estimates are initially biased an iterative process can be applied, identifying and removing outliers and repeating effect estimation until no outliers exceeding a given Q-statistic threshold are present. However, as in univariable MR, this is reliant upon such outliers being invalid. In cases where the majority of instruments are pleiotropic with a similar distribution of pleiotropic effects, it is possible that valid instruments will be identified as outliers. In these cases, the removal of outliers can lead to estimates converging towards biased estimates of 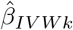.

### The RMVMR R package

The RMVMR R package is a tool designed to facilitate the implementation and visualisation of RMVMR analyses. RMVMR analyses should ideally be performed in five stages. First, summary GWAS data need to be obtained for a set of instruments, including instrument-exposure associations for all included exposures, instrument-outcome associations, and corresponding standard errors. With this complete, the data are formatted for downstream analyses using the format_rmvmr() function, and conditional instrument strength is evaluated using the strength_rmvmr() function. Causal effect estimates and tests for pleiotropic instruments can then be performed using the ivw_rmvmr() and pleiotropy_rmvmr() functions. Finally, the plot_rmvmr() function can be used to construct RMVMR plots. A flow chart showing each step for applying RMVMR using the RMVMR software package is provided in the supplementary material.

Outliers are detected based on their contribution to heterogeneity after adjustment, and are calculated with respect to each individual exposure. The significance level for identifying outliers can be defined by the user, and a data frame containing the Q-statistics for each individual variant is provided as an output from the pleiotropy_rmvmr() function. Identified outliers can then be followed up using external sources such as PhenoScanner or the MR Base online software platform [21, 22]. The RMVMR package builds upon the RadialMR and MVMR R packages, and can be used in conjunction with data obtained using the MR Base platform. Further details for the RMVMR software and installation instructions are available at https://github.com/WSpiller/RMVMR.

## Results

### Demonstrating the implementation of RMVMR through simulation

To demonstrate the implementation and advantages of RMVMR a simulation study is presented comprised of two components (simulations 1 and 2). In simulation 1 a single data frame is generated and analysed. This serves to illustrate how RMVMR analyses are implemented and interpreted in an individual case, including outlier detection and plot construction. Simulation 2 considers estimates of causal effect obtained from 1,000 data frames, highlighting broader features of RMVMR. Code for replicating the analyses is provided in the supplementary material, and the data set used in simulation 1 is the last of 1,000 generated for simulation 2. All RMVMR analyses are performed using the RMVMR R package.

Each data frame is simulated so as to include *N* = 200, 000 observations of *J* = 240 instruments *G*_*j*_, three exposures *X*_1−3_, a single unmeasured confounder *U* and an outcome *Y*. The data were generated using a data generating model conforming to equations (9 and 10). The set of instruments were generated so as to represent eight equal groups based on their association with one or more exposures, and were assigned arbitrary identification (rsid) numbers. Simulated groups of instruments include:

- Instruments associated with *X*_1_ only (group 1: rs1-30)
- Instruments associated with *X*_2_ only (group 2: rs31-60)
- Instruments associated with *X*_3_ only (group 3: rs61-90)
- Instruments associated with *X*_1_ and *X*_2_ (group 4: rs91-120)
- Instruments associated with *X*_1_ and *X*_3_ (group 5: rs121-150)
- Instruments associated with *X*_2_ and *X*_3_ (group 6: rs151-180)
- Instruments associated with *X*_1_, *X*_2_ and *X*_3_ (group 7: rs181-210)
- Instruments associated with *X*_1_, *X*_2_ and *X*_3_ with a direct effect on Y (group 8: rs211-240)

Non-zero associations between instruments and exposures were randomly sampled from a normal distribution with mean 0 and standard deviation 10. To ensure strong instruments were used in the analysis, values with an absolute value less than 2 were resampled. The effects of *X*_1_, *X*_2_ and *X*_3_ upon *Y* were defined as *β*_1_ = 1, *β*_2_ = 0.2, and *β*_3_ = − 0.5 respectively. Exposures were also simulated so as to be correlated, with a correlation coefficient ranging from − 0.5 to 0.5.

Instrument group 8 was subdivided into three equal groups, wherein instruments with a direct effect on *Y* are associated with one of the three exposures *X*_1−3_. The direct effects of instruments in group 8 were sampled from a normal distribution with mean 10 and standard deviation 5, resampling to ensure parameters had an absolute value greater than 2. Finally, each set of instrument-exposure estimates, as well as instrument-outcome associations, were obtained from separate non-overlapping samples.

#### Simulation 1

*Demonstrating the application of RMVMR using a single data frame*

Initially, univariable radial IVW models are applied using only instruments robustly associated with each exposure (F-statistic > 10). Radial MR estimates for each individual exposure are presented in Table 1, while univariable radial plots are provided in the supplementary material.

**Table 1.** Causal effect estimates^1^ obtained using radial MR and RMVMR models with differing exposure combinations^2^.

| Model | Estimate (se) | p-value | Q-statistic (p-value) |
| --- | --- | --- | --- |
| Univariable Radial IVW |  |  |  |
| $X_1$ | 1.051 (0.041) | $< 0.001$ | 9242.04 ($< 0.001$) |
| $X_2$ | 0.697 (0.069) | $< 0.001$ | 24012.2 ($< 0.001$) |
| $X_3$ | 0.342 (0.076) | $< 0.001$ | 29652.94 ($< 0.001$) |
| RMVMR ($X_1, X_2$) | | | |
| $X_1$ | 0.971 (0.045) | $< 0.001$ | 623.81 ($< 0.001$) |
| $X_2$ | 0.142 (0.046) | 0.002 | 621.41 ($< 0.001$) |
| RMVMR ($X_1, X_2, X_3$) | | | |
| $X_1$ | 1.098 (0.039) | $< 0.001$ | 544.08 ($< 0.001$) |
| $X_2$ | 0.311 (0.041) | $< 0.001$ | 554.90 ($< 0.001$) |
| $X_3$ | -0.422 (0.040) | $< 0.001$ | 557.61 ($< 0.001$) |
| Pruned RMVMR ($X_1, X_2, X_3$) | | | |
| $X_1$ | 1.001 (0.010) | $< 0.001$ | 23.66 ($> 0.999$) |
| $X_2$ | 0.201 (0.011) | $< 0.001$ | 26.46 ($> 0.999$) |
| $X_3$ | -0.497 (0.011) | $< 0.001$ | 22.90 ($> 0.999$) |

<sup>1</sup> True effects of each exposure: $\beta_1 = 1$ , $\beta_2 = 0.2$ , $\beta_3 = -0.5$ .
<sup>2</sup> Sample size $N = 200,000$

In Table 1 we can see that effect estimates exhibit substantial bias when estimated using univariable radial IVW. The high Q-statistics estimated for each exposure provide evidence of heterogeneity in estimates obtained using each instrument individually, suggesting potential violations of assumption IV3. When using a two exposure RMVMR model including exposures *X*_1_ and *X*_2_, there continues to be evidence of bias. In this case the observed bias is smaller in magnitude, reflecting how adjustment for both *X*_1_ and *X*_2_ accounts for a proportion of the pleiotropic bias observed in univariable analyses. This is expected given instrument groups 4, 7, and 8 are simultaneously associated with exposures *X*_1_ and *X*_2_. This interpretation is further supported by a notable decrease in observed heterogeneity for each exposure. For reference, the estimated conditional F-statistics for *X*_1_ and *X*_2_ were 111.32 and 112.22 respectively.

The inclusion of exposure *X*_3_ adjusts for associations between instruments and the outcome through *X*_3_ which are not mediated downstream by either *X*_1_ or *X*_2_. This again results in a substantial decrease in heterogeneity, although the continued presence of instruments from group 8 has the effect of inducing pleiotropic bias. The conditional F-statistics for each exposure were 105.94, 98.21, and 103.46 respectively.

By removing instruments identified as outliers on the basis of their contribution to global heterogeneity, it is possible to perform a pruned analysis using the iterative approach previously described. Calculating the individual Q-statistic for each instrument with respect to each exposure, a total of 16 SNPs are identified as outliers using a p-value threshold of 0.05, shown in Figure 5. From Figure 5 it can be seen that all identified outliers correspond to group 8 (rs211-240); instruments generated so as to violate assumption MVMR3 by having a direct effect on the outcome *Y*. Consequently, removing these instruments will have the effect of reducing pleiotropic bias in RMVMR analyses. In Table 1 estimates obtained using the pruned RMVMR approach show no evidence of bias or substantial heterogeneity. The conditional F-statistics for the pruned analysis were 115.56, 107.42, and 114.11 for exposures *X*_1_, *X*_2_, and *X*_3_.

**Fig 5.**
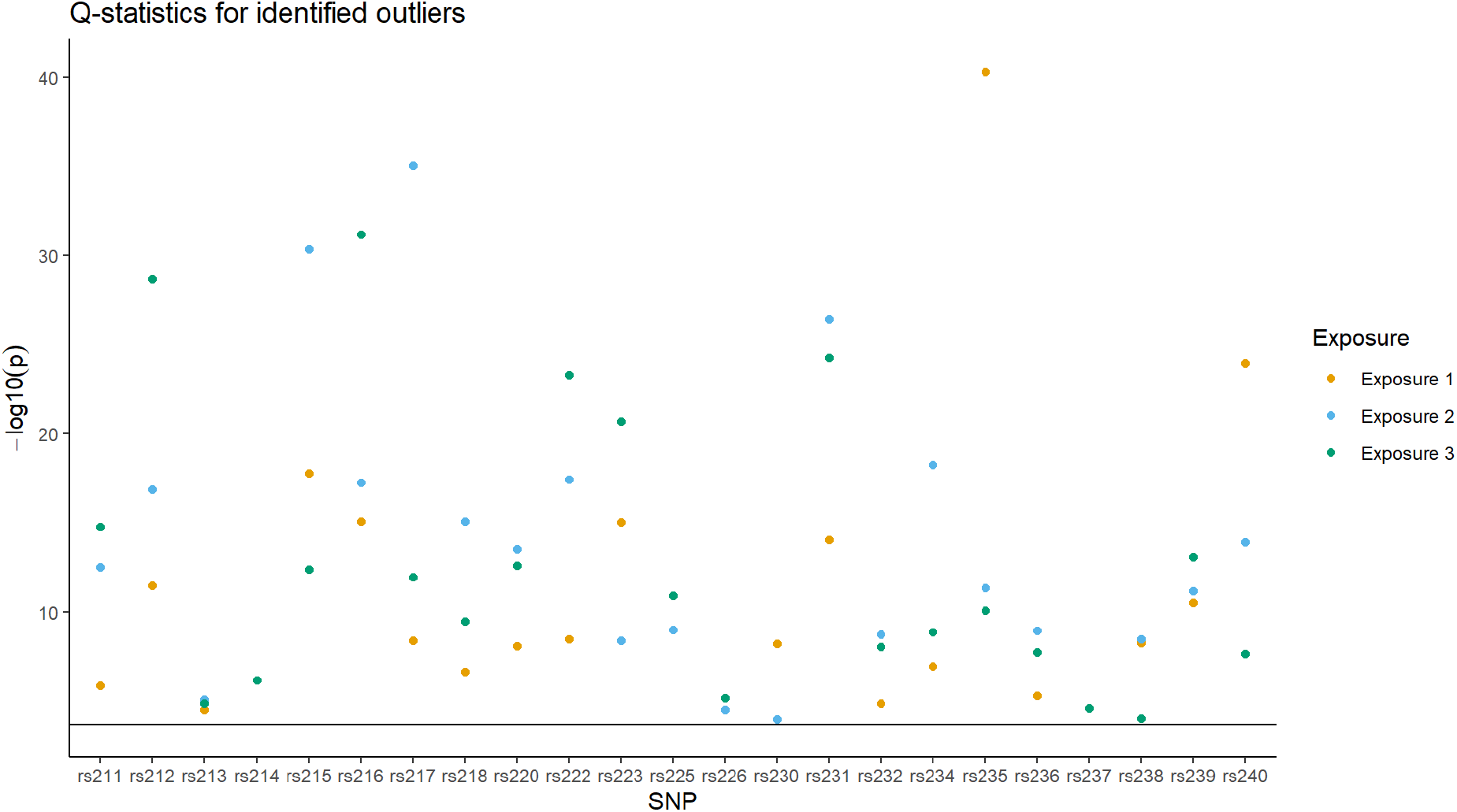
Scatter plot showing instruments which are identified as outliers using the p-value for their contribution to observed heterogeneity. A dotted line is shown representing the p-value threshold for identifying outliers (*p <* 0.05). All instruments correspond to Group 8 (rs211-240) for which a directional pleiotropic effect is present.

Fig 6 shows RMVMR plots corresponding to the models adopted in simulation 1. In Fig 6s observations do not appear to converge towards their respective effect estimates, instead forming two widely dispersed clusters. The extent to which observations diverge from their corresponding direct effect estimate is representative to their contribution to global heterogeneity, and consequently serves as an indicator of MVMR3 violation.

**Fig 6.**
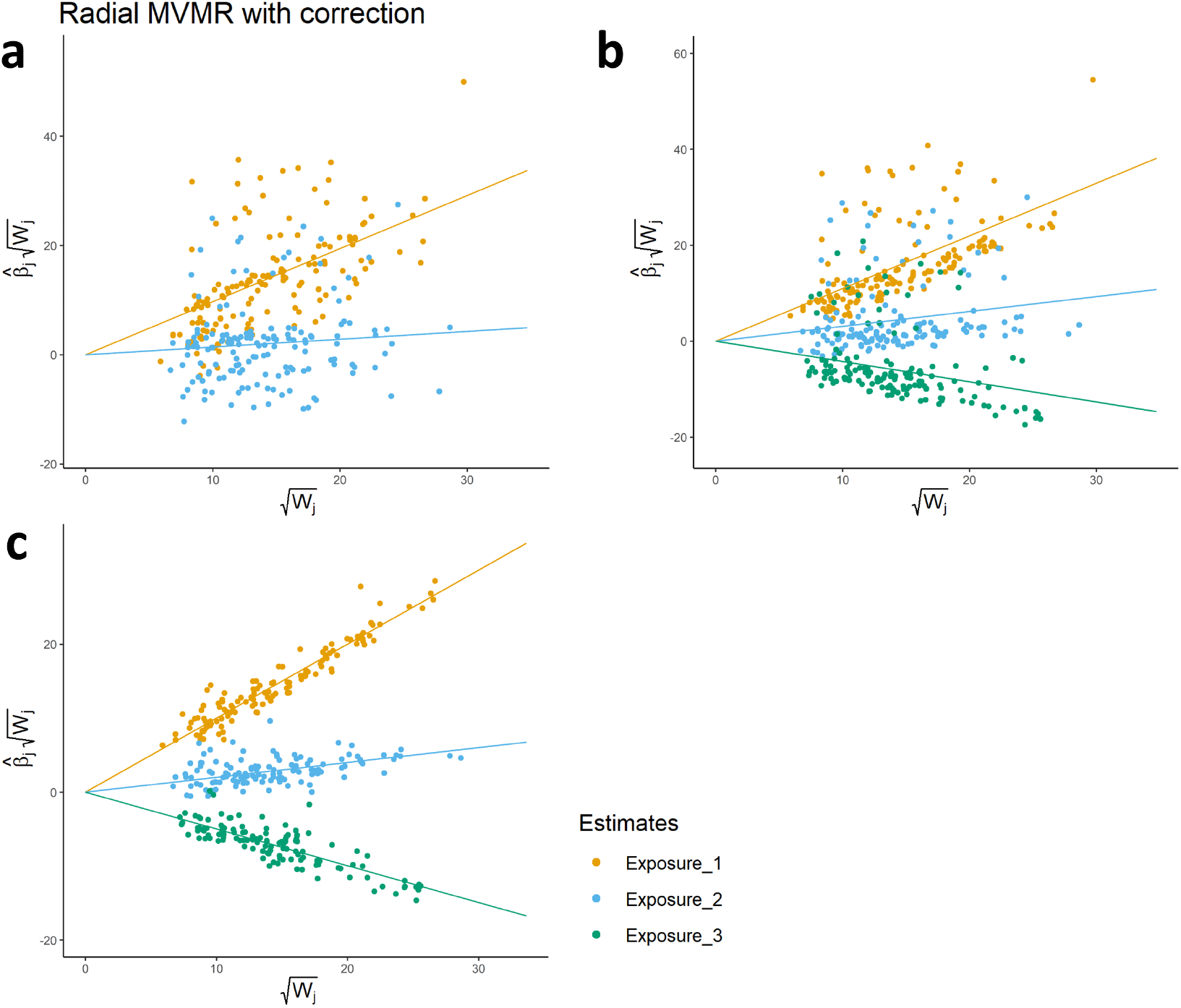
Panel showing radial MVMR plots corresponding to each of the simulated analyses presented in Table 1. Fig 6a represents the two-exposure model, while Fig 6b includes all measured exposures. Fig 6c, shows a plot generated after pruning pleiotropic SNPs identified in Fig 5.

In Fig 6b the inclusion of exposure *X*_3_ has the effect of substantially reducing global heterogeneity. In this case, a pattern emerges where it is possible to visually identify valid instruments defined in the simulation. The inclusion of instruments from group 8, however, induces pleiotropic bias in causal effects. The continued presence of pleiotropic instruments is indicated by the continued presence of substantial heterogeneity.

Following the systematic pruning of outliers, Fig 6c shows how remaining instruments converge towards unbiased estimates of direct effect for each exposure. Notably, the change of scale on the y-axis reflects the reduction in relative distance from each observation to their respective superimposed regression line, and the absence of substantial heterogeneity suggests an absence of pleiotropic bias. It is, however, important to once again emphasise that such an interpretation is predicated on MVMR3 violating instruments being identified as outliers, and care should be taken when considering outlier removal.

#### Simulation 2

*Evaluating the performance of RMVMR over multiple iterations*

In simulation 1 attention was given to the implementation and interpretation of RMVMR using a single data set. To provide a more concrete demonstration of how RMVMR can lead to an overall reduction in pleiotropic bias, we repeat the previous analyses using a total of 1,000 independent data sets. Mean effect estimates, standard errors, and F-statistics are presented in Table 2, and illustrated in Fig 7.

**Table 2.** Mean causal effect estimates^1^ obtained using RMVMR models across 1,000 independent data sets^2^) from simulation 2.

| Model | Mean estimate | Mean std.error | Mean F-statistic<sup>3</sup> |
| --- | --- | --- | --- |
| RMVMR ($X_1, X_2, X_3$) | | | |
| $X_1$ | 1.010 | 0.038 | 107.30 |
| $X_2$ | 0.311 | 0.041 | 98.12 |
| $X_3$ | -0.418 | 0.040 | 103.50 |
| Pruned RMVMR ($X_1, X_2, X_3$) | | | |
| $X_1$ | 0.999 | 0.010 | 117.01 |
| $X_2$ | 0.206 | 0.011 | 107.77 |
| $X_3$ | -0.486 | 0.010 | 115.24 |

<sup>1</sup> True effects of each exposure: $\beta_1 = 1$ , $\beta_2 = 0.2$ , $\beta_3 = -0.5$ .
<sup>2</sup> Sample size $N = 200,000$
<sup>3</sup> Conditional F-statistic

**Fig 7.**
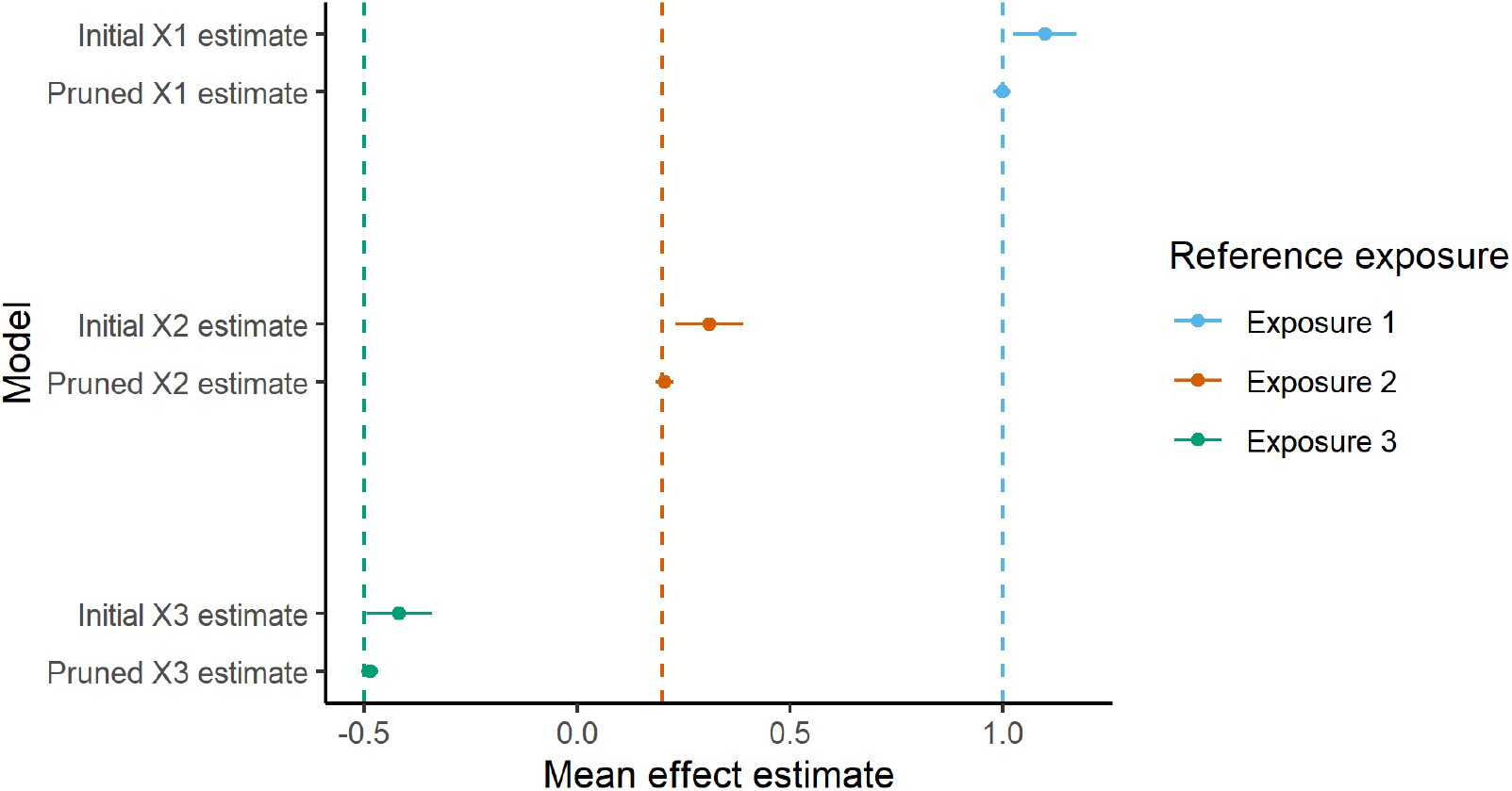
Forest plot showing mean effect estimates and 95% confidence intervals for the effect of exposures *X*_1_, *X*_2_, and *X*_3_ following simulation 2. Vertical dashed lines depict defined true effects, coloured by exposure. Initial estimates indicate RMVMR estimates obtained prior to outlier pruning.

When evaluating the performance of RMVMR across multiple iterations, we can see that identifying and pruning outliers in this case consistently leads to a reduction in pleiotropic bias. It is also interesting to highlight that conditional instrument strength for each exposure is greater after removing outlying instruments. This is a consequence of removing noise from instruments in group 8, such that the reduction in heterogeneity affords more accurate prediction of each exposure using the remaining instruments.

When evaluating the performance of RMVMR across multiple iterations, we can see that identifying and pruning outliers in this case consistently leads to a reduction in pleiotropic bias, as well as a substantial increase in estimate precision. It is also interesting to highlight that conditional instrument strength for each exposure is greater after removing outlying instruments. This is a consequence of removing noise induced by instruments in group 8, such that the reduction in heterogeneity affords more accurate prediction of each exposure using the remaining instruments.

#### Applied analysis: Lipid fractions and coronary heart disease

To demonstrate the RMVMR approach in an applied setting we consider the effects of lipid fractions, specifically HDL, LDL, and triglycerides, upon CHD. SNP-exposure estimates were obtained from previously published GWAS summary data, using data from the Global Lipids Genetics Consortium presented in Willer et al [19]. Each lipid fraction was recorded in mg/Dl, and standardised before GWAS were performed. SNP-outcome associations were obtained from the CARDIoGRAMplusC4D Consortium as presented in Nikpay et al [18]. CHD associations are presented on a log-odds scale, and were obtained using logistic regression. All SNP associations were obtained using the MRBase online platform [21]. For each exposure, univariable radial MR analyses are performed, after which radial MVMR models including all exposures are fit to the data.

Univariable radial MR analyses were performed for HDL, LDL, and triglycerides, using only SNPs identified as robustly associated with each exposure (*p* < 5 × 10^−8^). Independent SNPs were selecting using a linkage disequilibrium clumping threshold of *R*^2^ < 0.001, and palindromic SNPs were also removed prior to performing analyses. This resulted in a total of 87 SNPs for HDL, 67 SNPs for LDL, and 40 SNPs for triglycerides being selected for subsequent analyses. Effect estimates in Table 3 have been transformed so as to be interpreted on an odds ratio scale. Mean F-statistics for SNPs used in univariable analyses are presented in Table 3, and corresponding plots for univariable analyses are provided in the supplementary material.

**Table 3.** Causal effect estimates obtained using radial MR and radial MVMR models, estimating the effect of lipid fractions (HDL, LDL, and triglycerides) on CHD.

| Model | Estimate (se) | p-value | Q-statistic (p-value) | F-Statistic |
| --- | --- | --- | --- | --- |
| Univariable Radial IVW |  |  |  |  |
| HDL<sup>1</sup> | 0.829 (0.058) | 0.001 | 444.90 (< 0.001) | 121.6 |
| LDL<sup>1</sup> | 1.499 (0.055) | < 0.001 | 229.35 (< 0.001) | 100.1 |
| Triglycerides<sup>1</sup> | 1.255 (0.062) | < 0.001 | 162.81 (< 0.001) | 170.3 |
| RMVMR |  |  |  |  |
| HDL | 0.899 (0.053) | 0.046 | 144.24 (0.004)<sup>2</sup> | 42.9 |
| LDL | 1.408 (0.057) | < 0.001 | 109.36 (0.020)<sup>2</sup> | 39.2 |
| Triglycerides | 1.123 (0.064) | 0.074 | 84.13 (0.055)<sup>2</sup> | 29.1 |
| Pruned RMVMR |  |  |  |  |
| HDL | 0.939 (0.042) | 0.135 | 74.39 (0.910)<sup>2</sup> | 43.6 |
| LDL | 1.399 (0.050) | < 0.001 | 40.13 (0.997)<sup>2</sup> | 32.2 |
| Triglycerides | 1.146 (0.055) | 0.014 | 46.16 (0.869)<sup>2</sup> | 24.9 |

<sup>1</sup> Estimates obtained using univariable radial MR.
<sup>2</sup> Calculated using corrected ratio estimates

Considering the univariable radial IVW analyses there appears to be evidence of a positive association of LDL and triglycerides with CHD, in contrast to HDL which shows evidence of a protective effect. However, evaluating heterogeneity across the range of individual ratio estimates for each exposure indicates that one or more SNPs may be pleiotropic, potentially violating assumption IV3. This is indicated by high Q-statistics for each exposure, as seen in Table 3.

Previous research has highlighted how individual SNPs simultaneously associated with multiple lipid fractions, coupled with an observed strong correlation between phenotypes, can result in pleiotropic bias. To account for such relationships, it is possible to fit a radial MVMR model incorporating each exposure simultaneously. Radial MVMR estimates for each exposure are shown in Table 3, showing the effect of each lipid fraction to be directionally consistent, though smaller in magnitude, compared to univariable estimates. Assuming that the observed heterogeneity in univariable analyses is the result of omitting one or more lipid fractions, performing a correction for each observation and re-evaluating heterogeneity using radial MVMR should show evidence of a substantial Q-statistic decrease. Table 3 shows a substantial decrease in observed heterogeneity when fitting the radial MVMR model, though HDL and LDL still show evidence of significant global heterogeneity using a Q-statistic value threshold of *p* < 0.05.

As in the previous simulation analyses, it is possible to identify and remove SNPs which contribute a substantial degree of heterogeneity within analyses. For this analysis, SNPs with a significantly high Q-statistic for either HDL, LDL, or triglycerides were omitted, subsequently estimating causal effects through an iterative process until no outliers were detected. These results are shown in Table 3, where effect estimates remain directionally consistent with the initial radial MVMR analysis. Performing the pruned analysis results in effects of smaller magnitude for HDL and LDL, and the effects of all exposures are estimated with greater precision. Importantly, the conditional F-statistic for each exposure remains at a similar level to the initial radial MVMR analysis, limiting the extent to which differences in estimation are the result of bias due to weak instruments being used after pruning.

Removing SNPs which exhibit heterogeneity does not necessarily imply that estimates will be less biased. If the majority of SNPs exhibit pleiotropic effects in a similar direction and magnitude, it is possible that SNPs satisfying the MVMR assumptions will be removed. To consider this possibility it is important to follow-up identified outliers using external data, focusing on associations with phenotypes for which a pleiotropic association is plausible. In the pruned analysis, a total of 17 SNPs were identified and removed as outliers (see supplementary material). Using the PhenoScanner online platform to evaluate potential pleiotropic pathways, there did not appear to be a consistent pattern across the set of removed SNPs, though phenotypes such as diastolic blood pressure are present [22].

The radial MVMR estimates are visualised in Figure 8, and adjusted Q-statistics are presented in Table 3. The reduction in observed heterogeneity suggests that univariable analyses exhibit bias when failing to account for pleiotropic associations through other lipid fractions. A multivariable model would therefore appear to be a more effective approach in this instance. The plots shown in Figure 8a and 8b show the estimates obtained without pruning SNPs based on their heterogeneity contribution, while plots 8c and 8d show the plots constructed after removing observed outliers. In this case, the reduction in global heterogeneity is clear and primarily reflected by the change of scale on the y-axis, in combination with the data points for each exposure being substantially closer to their corresponding superimposed regression lines.

**Fig 8.**
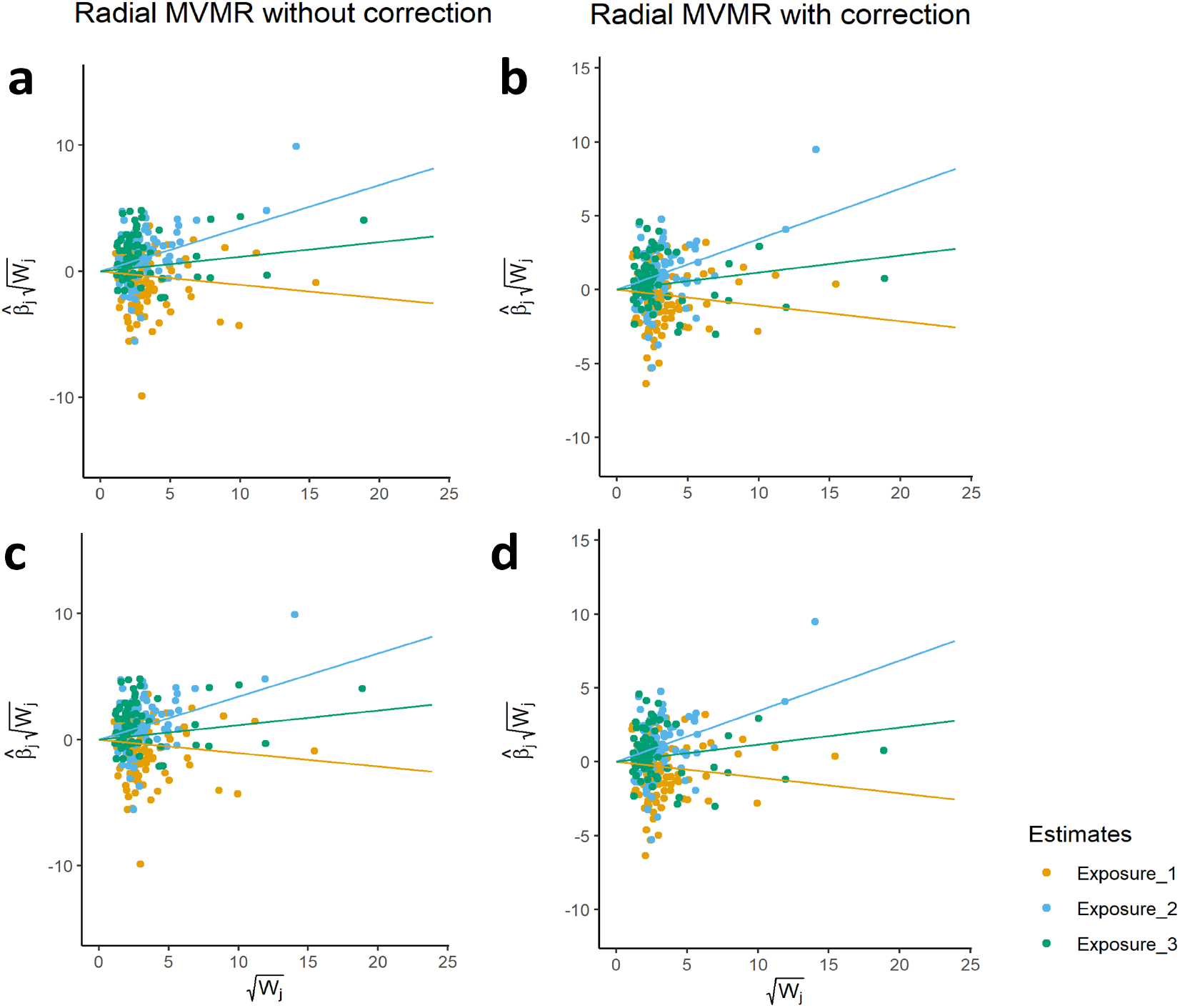
Panel showing RMVMR plots for applied analysis using HDL, LDL, and triglycerides. Observations correspond to ratio estimates and weightings with respect to each exposure. Regression lines represent MVMR causal effect estimates coloured by exposure. Fig 8a-8b correspond to the radial MVMR estimates prior to performing heterogeneity pruning, while Fig 8c-8d are constructed using heterogeneity pruned summary data.

When pruning for outliers the effect of HDL is greatly attenuated, showing no evidence of an effect on CHD. LDL and triglycerides continue to show evidence of a positive association, with LDL having the most substantial impact on CHD risk. Provided a majority of valid SNPs with respect to their weighting have been used, this would suggest that LDL and triglycerides represent promising targets for CHD prevention.

## Discussion

Radial MR and MVMR approaches facilitate the assessment of pleiotropic associations between genetic variants and phenotypes using GWAS summary data. Conducting analyses within a radial framework allows for outliers to be effectively visualised, and MVMR analyses allow for the direct effects of multiple exposures to be estimated simultaneously. Radial MVMR builds upon both these existing methods, providing a means for visualising MVMR approaches absent until this point, and justifying the use of MVMR where relevant exposure data are available. We propose that the radial MVMR approach be used to assist in communicating key findings as a visual aid, and also as a sensitivity analysis for identifying pleiotropic bias using adjusted heterogeneity statistics.

Radial MVMR builds of existing work which leverages publicly available genetic data to correct for pleiotropic bias. The work in this paper differs in providing further diagnostic tools, as well as a means of visualising MVMR analyses. In this way, it is potentially easier to identify specific SNPs which may serve as outliers in an analyses, warranting subsequent follow up.

When implementing the radial MVMR approach it is crucial to consider the underlying assumptions of MVMR. Instruments selected should be sufficiently strong so as to overcome substantial weak instrument bias, estimated using the conditional F-statistic. Specific to radial MVMR, the correction of individual ratio estimates is reliant upon unbiased estimates of the direct effect of each exposure. As demonstrated in the simulation study, in cases where direct effects exhibit biases the subsequent correction for each SNP will be incorrect. However, this is unlikely to result in a substantial reduction in heterogeneity unless the distribution of pleiotropic associations is similar, and will likely still indicate that pleiotropic bias may be present. This is due to the differing contribution of each SNP towards heterogeneity, as a consequence of differences in their relationship within one or more pleiotropic pathways.

As previously emphasised, care should be taken to consider identified outliers and phenotypic associations which could plausibly form horizontal pleiotropic pathways. If a majority of instruments have direct effects upon an outcome, and the distribution of such direct effects is similar, it is likely that SNPs satisfying the MVMR assumptions will be identified as outliers. As a consequence, the removal of such SNPs would result in estimates converging toward the biased estimate produced by such pleiotropic SNPs. Decisions to down weight or remove outliers during an analysis should be made with consideration of the biological mechanisms underlying observed SNP-phenotype associations, and adequate justification. It is with this in mind that a general heterogeneity pruning function has not been incorporated within the RMVMR R package, though code for performing such analyses is provided in the supplementary material and is available at https://github.com/WSpiller/RMVMR_Analyses.

A further issue related to the applied analysis is the use of binary outcomes in summary MR analyses. When using SNP-outcome associations estimated on a log-odds scale, it is possible that causal estimates will be correlated with their precision, introducing heterogeneity which is not a consequence of pleiotropic associations [6, 23]. This issue, which is a wider issue within the summary MR literature, warrants careful consideration prior to performing analyses, and care should be taken in evaluating as an indicator of pleiotropy when a binary outcome is used [23].

Finally, it should be noted that while first-order weights have been used throughout this paper, radial approaches allow for a wide-range of weighting options to be used. As arbitrary weights can be used, should be possible for modified second order weights to be incorporated with a radial MVMR model. Such weights may prove more effective than first-order weights, as they incorporate the precision of SNP-exposure estimates mitigating violations of the NO-Measurement Error (NOME) assumption in summary MR [5]. Future work will explore how differing weight specifications can improve estimation using radial MVMR.

## Data Availability

This study only uses simulated data and publicly available summary data from genome-wide association studies. Scripts for replicating the analyses and summary GWAS data used for applied analyses are provided at https://github.com/WSpiller/RMVMR_Analyses.

https://github.com/WSpiller/RMVMR

## Supporting information

### Data availability

Scripts for reproducing the simulated examples in this paper, in addition to summary GWAS data for applied analyses are available at https://github.com/WSpiller/RMVMR_Analyses. Summary GWAS data for applied analyses is publicly available, and was obtained from the OpenGWAS catalogue (https://gwas.mrcieu.ac.uk/).

### Contributions

WS, JB, and ES contributed to the idea and development of the RMVMR method. WS performed the analyses and created the RMVMR R package. WS drafted the manuscript, and JB and ES provided critical input throughout the drafting process.

## Acknowledgments

The authors would like to thank Dr Tom Palmer for his help in improving and maintaining the RMVMR R package.

