## Supplementary material for "Estimating and visualising multivariable Mendelian randomization analyses within a radial framework": Code,data, and additional information: 1_Data_gen.R

```
##############################################
###   Install and load relevant packages   ###
##############################################


#install.packages("remotes")
#library(remotes)
#install_github("WSpiller/RMVMR", build_opts = c("--no-resave-data", "--no-manual"), build_vignettes = TRUE)
#install_github("WSpiller/MVMR", build_opts = c("--no-resave-data", "--no-manual"), build_vignettes = TRUE)
#install_github("WSpiller/RadialMR", build_opts = c("--no-resave-data", "--no-manual"), build_vignettes = TRUE)

#Set random gen.seed
set.seed(52431)

#Load libraries into R
library(RMVMR)
library(RadialMR)
library(MVMR)
library(ggplot2)

#Set number of iterations (For single iteration selecting 100th simulation for simulation 1)
K<-100

#################################
### Set simulation parameters ###
#################################

# G-X Parameters

X1gammas<-rnorm(30,0,5)
X1gammas<-c(X1gammas,rep(0,30))
X1gammas<-c(X1gammas,rep(0,30))
X1gammas<-c(X1gammas,rnorm(30,0,5))
X1gammas<-c(X1gammas,rnorm(30,0,5))
X1gammas<-c(X1gammas,rep(0,30))
X1gammas<-c(X1gammas,rnorm(30,0,5))
X1gammas<-c(X1gammas,rnorm(30,0,5))

X1gammas<-abs(X1gammas)

for(i in 1:240){
  X1gammas[i]<-X1gammas[i] + sign(X1gammas[i]) * 5
}

X2gammas<-rep(0,30)
X2gammas<-c(X2gammas,rnorm(30,0,5))
X2gammas<-c(X2gammas,rep(0,30))
X2gammas<-c(X2gammas,rnorm(30,0,5))
X2gammas<-c(X2gammas,rep(0,30))
X2gammas<-c(X2gammas,rnorm(30,0,5))
X2gammas<-c(X2gammas,rnorm(30,0,5))
X2gammas<-c(X2gammas,rnorm(30,0,5))

X2gammas<-abs(X2gammas)

for(i in 1:240){
  X2gammas[i]<-X2gammas[i] + sign(X2gammas[i]) * 5
}

X3gammas<-rep(0,30)
X3gammas<-c(X3gammas,rep(0,30))
X3gammas<-c(X3gammas,rnorm(30,0,5))
X3gammas<-c(X3gammas,rep(0,30))
X3gammas<-c(X3gammas,rnorm(30,0,5))
X3gammas<-c(X3gammas,rnorm(30,0,5))
X3gammas<-c(X3gammas,rnorm(30,0,5))
X3gammas<-c(X3gammas,rnorm(30,0,5))

X3gammas<-abs(X3gammas)

for(i in 1:240){
  X3gammas[i]<-X3gammas[i] + sign(X3gammas[i]) * 5
}

#Define effect of X1
beta1<-1

#Define effect of X2
beta2<-0.2

#Define effect of X3
beta3<-(-0.5)

# G-Y Parameters

alphavec<-rep(0,210)
alphavec<-abs(c(alphavec,rnorm(30,10,5)))

for(i in 1:240){
  alphavec[i]<-alphavec[i] + sign(alphavec[i]) * 2
}

#Define empty vectors for estimates and standard errors without pruning
X1est<-rep(0,K)
X2est<-rep(0,K)
X3est<-rep(0,K)
X1se<-rep(0,K)
X2se<-rep(0,K)
X3se<-rep(0,K)

#Define empty vectors for estimates and standard errors after pruning
X1pest<-rep(0,K)
X2pest<-rep(0,K)
X3pest<-rep(0,K)
X1pse<-rep(0,K)
X2pse<-rep(0,K)
X3pse<-rep(0,K)

#Define empty vectors for conditional f-statistics without pruning
X1f<-rep(0,K)
X2f<-rep(0,K)
X3f<-rep(0,K)

#Define empty vectors for conditional f-statistics after pruning
X1fp<-rep(0,K)
X2fp<-rep(0,K)
X3fp<-rep(0,K)

#########################
###   Generate data   ###
#########################

for(k in 1:K){
  
  #Print iteration number
  print(k)
  
  #Set number of instruments to 240
  
  J<-240
  
  #Set sample size
  N<-200000
  
  #Generate set of instruments
  G.dat<-data.frame(rep(0,N))
  
  for(i in 1:J){
    G.dat[,i]<-rnorm(N,0,1)
  }
  
  #Generate unmeasured confounder
  U<-rnorm(N,0,1)
  
  #  #shared effect between exposures
  X1X2<-rnorm(N,0,1)
  X1X3<-rnorm(N,0,1)
  X2X1<-rnorm(N,0,1)
  X2X3<-rnorm(N,0,1)
  X3X1<-rnorm(N,0,1)
  X3X2<-rnorm(N,0,1)
  
  #Generate exposures
  X1<-1 + 1*U + rnorm(N,0,1) + rnorm(1,0,5)*X1X2 + rnorm(1,0,5)*X1X3 + rnorm(1,0,5)*X3X1 + rnorm(1,0,5)*X2X1
  X2<-1 + 1*U + rnorm(N,0,1) + rnorm(1,0,5)*X1X2 + rnorm(1,0,5)*X2X1 + rnorm(1,0,5)*X2X3 + rnorm(1,0,5)*X3X2
  X3<-1 + 1*U + rnorm(N,0,1) + rnorm(1,0,5)*X1X3 + rnorm(1,0,5)*X2X3 + rnorm(1,0,5)*X3X1 + rnorm(1,0,5)*X3X2
  
  
  #Generate outcome
  Y<-1 + rnorm(N,0,1)
  
  #Pleiotropic effects for instruments
  for (i in c(1:240)){
    Y<-Y + alphavec[i]*G.dat[,i]
  }
  
  #X1 associations
  for (i in 1:240){
    X1<-X1 + X1gammas[i]*G.dat[,i]
  }
  
  #X2 associations
  for (i in 1:240){
    X2<-X2 + X2gammas[i]*G.dat[,i]
  }
  
  #X3 associations
  for (i in 1:240){
    X3<-X3 + X3gammas[i]*G.dat[,i]
  }
  
  #Generate outcome
  Y<- Y + beta1*X1 + beta2*X2 + beta3*X3 + 1*U + rnorm(N,0,1)
  
  #Create combined dataframe
  c.dat<-data.frame(X1,X2,X3,Y)
  c.dat<-cbind(G.dat,c.dat)
  for(i in 1:J){
    names(c.dat)[i]<-paste("rs",i,sep="")
  }
  
  #Divide sample into non-overlapping subsets for generating exposure and outcome associations
  exposure.sampleX1<-c.dat[1:50000,]
  exposure.sampleX2<-c.dat[50001:100000,]
  exposure.sampleX3<-c.dat[100001:150000,]
  outcome.sampleY<-c.dat[150001:200000,]
  
  # Estimate G-X1 associations
  
  gammaX1hat<-rep(0,J)
  segammaX1hat<-rep(0,J)
  pgammaX1hat<-rep(0,J)
  
  for(i in 1:J){
    
    gammaX1hat[i]<-summary(lm(exposure.sampleX1[,J+1]~exposure.sampleX1[,i]))$coef[2,1]
    segammaX1hat[i]<-summary(lm(exposure.sampleX1[,J+1]~exposure.sampleX1[,i]))$coef[2,2]
    pgammaX1hat[i]<-summary(lm(exposure.sampleX1[,J+1]~exposure.sampleX1[,i]))$coef[2,4]
    
  }
  
  # Estimate G-X2 associations
  
  gammaX2hat<-rep(0,J)
  segammaX2hat<-rep(0,J)
  pgammaX2hat<-rep(0,J)
  
  for(i in 1:J){
    gammaX2hat[i]<-summary(lm(exposure.sampleX2[,J+2]~exposure.sampleX2[,i]))$coef[2,1]
    segammaX2hat[i]<-summary(lm(exposure.sampleX2[,J+2]~exposure.sampleX2[,i]))$coef[2,2]
    pgammaX2hat[i]<-summary(lm(exposure.sampleX2[,J+2]~exposure.sampleX2[,i]))$coef[2,4]
    
  }
  
  # Estimate G-X3 associations
  
  gammaX3hat<-rep(0,J)
  segammaX3hat<-rep(0,J)
  pgammaX3hat<-rep(0,J)
  
  for(i in 1:J){
    gammaX3hat[i]<-summary(lm(exposure.sampleX3[,J+3]~exposure.sampleX3[,i]))$coef[2,1]
    segammaX3hat[i]<-summary(lm(exposure.sampleX3[,J+3]~exposure.sampleX3[,i]))$coef[2,2]
    pgammaX3hat[i]<-summary(lm(exposure.sampleX3[,J+3]~exposure.sampleX3[,i]))$coef[2,4]
    
  }
  
  # Estimate G-Y associations
  
  gammaYhat<-rep(0,J)
  segammaYhat<-rep(0,J)
  for(i in 1:J){
    
    gammaYhat[i]<-summary(lm(outcome.sampleY[,J+4]~outcome.sampleY[,i]))$coef[2,1]
    segammaYhat[i]<-summary(lm(outcome.sampleY[,J+4]~outcome.sampleY[,i]))$coef[2,2]
    
  }
  
  #Create dataframe containing summary data
  
  sum.data<-data.frame(names(c.dat[1:J]),gammaX1hat,gammaX2hat,gammaX3hat,
                       segammaX1hat,segammaX2hat,segammaX3hat,
                       gammaYhat,segammaYhat,pgammaX1hat,pgammaX2hat,pgammaX3hat)
  
  names(sum.data)[1]<-"SNP"
  
  
  ################################################
  ###   Radial MVMR Analyses: X1, X2, and X3   ###
  ################################################
  
  # Select instruments for any exposure
  
  X1X2X3datuni<-sum.data
  
  #Format the X1-X2 data
  X1X2X3f.data<-format_rmvmr(sum.data[,2:4], sum.data[,8], sum.data[,5:7], sum.data[,9], sum.data[,1])
  
  #Estimate conditional instrument strength
  strength_X1X2X3<- suppressWarnings(strength_mvmr(X1X2X3f.data))
  
  #Save conditional F statistic for exposure 1
  X1f[k]<-strength_X1X2X3[1]
  
  #Save conditional F statistic for exposure 2
  X2f[k]<-strength_X1X2X3[2]
  
  #Save conditional F statistic for exposure 3
  X3f[k]<-strength_X1X2X3[3]
  
  #Estimate causal effects  using Radial MVMR
  X1X2X3res<-ivw_rmvmr(X1X2X3f.data,F)
  
  #Save effect estimate and se for exposure 1
  X1est[k]<-X1X2X3res$coef[1,1]
  X1se[k]<-X1X2X3res$coef[1,2]
  
  #Save effect estimate and se for exposure 2
  X2est[k]<-X1X2X3res$coef[2,1]
  X2se[k]<-X1X2X3res$coef[2,2]
  
  #Save effect estimate and se for exposure 3
  X3est[k]<-X1X2X3res$coef[3,1]
  X3se[k]<-X1X2X3res$coef[3,2]
  
  #Estimate pleiotropic effects and detect outliers
  pleioX1X2X3<-pleiotropy_rmvmr(X1X2X3f.data,X1X2X3res)
  
  #############################################################
  ###   Radial MVMR Analyses: X1, X2, and X3 with pruning   ###
  #############################################################
  X1X2X3fp.data<-X1X2X3f.data
  
  outliers<-"start"
  
  while(!is.null(outliers)){
    
    X1X2X3pres<-ivw_rmvmr(X1X2X3fp.data,F)
    pleioX1X2X3p<-pleiotropy_rmvmr(X1X2X3fp.data,X1X2X3pres)
    
    ###Prune###
    
    if(min(pleioX1X2X3p$qdat$qj_p)< 0.05){
      outliers<-pleioX1X2X3p$qdat[pleioX1X2X3p$qdat$qj_p <0.05,]$snp
      outliers<-unique(outliers)
      
      X1X2X3fp.data<-X1X2X3fp.data[!X1X2X3fp.data$SNP %in% outliers,]
      
    }else{
      outliers<-NULL
      
    }
    
  }
  
  #Estimate conditional instrument strength
  strength_X1X2X3p<- suppressWarnings(strength_mvmr(X1X2X3fp.data))
  
  #Save conditional F statistic for exposure 1
  X1fp[k]<-strength_X1X2X3p[1]
  
  #Save conditional F statistic for exposure 2
  X2fp[k]<-strength_X1X2X3p[2]
  
  #Save conditional F statistic for exposure 3
  X3fp[k]<-strength_X1X2X3p[3]
  
  #Estimate causal effects  using Radial MVMR
  X1X2X3resp<-ivw_rmvmr(X1X2X3fp.data,F)
  
  #Save effect estimate and se for exposure 1
  X1pest[k]<-X1X2X3resp$coef[1,1]
  X1pse[k]<-X1X2X3resp$coef[1,2]
  
  #Save effect estimate and se for exposure 2
  X2pest[k]<-X1X2X3resp$coef[2,1]
  X2pse[k]<-X1X2X3resp$coef[2,2]
  
  #Save effect estimate and se for exposure 3
  X3pest[k]<-X1X2X3resp$coef[3,1]
  X3pse[k]<-X1X2X3resp$coef[3,2]
  
}
```
