## Supplementary material for "Estimating and visualising multivariable Mendelian randomization analyses within a radial framework": Code,data, and additional information: 2_Simulation1.R

```
##############################################
###   Install and load relevant packages   ###
##############################################


#install.packages("remotes")
#library(remotes)
#install_github("WSpiller/RMVMR", build_opts = c("--no-resave-data", "--no-manual"), build_vignettes = TRUE)
#install_github("WSpiller/MVMR", build_opts = c("--no-resave-data", "--no-manual"), build_vignettes = TRUE)
#install_github("WSpiller/RadialMR", build_opts = c("--no-resave-data", "--no-manual"), build_vignettes = TRUE)

#Set random gen.seed
set.seed(52431)

#Load libraries into R
library(RMVMR)
library(RadialMR)
library(MVMR)
library(ggplot2)

#Load data to obtain 100th iteration for simulation 1
load("sim1.RData")

#############################
###  Sim 1 plots   ##########
#############################

#Select only instruments for exposure 1
X1datuni<-sum.data[sum.data$pgammaX1hat<0.05,]

#Format exposure 1 data
X1f.data<-format_radial(X1datuni[,2], X1datuni[,8], X1datuni[,5], X1datuni[,9], X1datuni[,1])

#Estimate causal effect of exposure 1 on Y
resX1uni<-ivw_radial(X1f.data,0.05,1,0.0001,T)

#Generate univariable radial plot for exposure 1
plotX1<-plot_radial(resX1uni,F,F,F)

png("Sim11AX1R.png",width=7,height=5.25,units="in",res=500)
plotX1
dev.off()

#Select only instruments for exposure 1
X2datuni<-sum.data[sum.data$pgammaX2hat<0.05,]

#Format exposure 1 data
X2f.data<-format_radial(X2datuni[,3], X2datuni[,8], X2datuni[,6], X2datuni[,9], X2datuni[,1])

#Estimate causal effect of exposure 1 on Y
resX2uni<-ivw_radial(X2f.data,0.05,1,0.0001,T)

#Generate univariable radial plot for exposure 1
plotX2<-plot_radial(resX2uni,F,F,F)

png("Sim11AX2R.png",width=7,height=5.25,units="in",res=500)
plotX2
dev.off()

#Select only instruments for exposure 1
X3datuni<-sum.data[sum.data$pgammaX3hat<0.05,]

#Format exposure 1 data
X3f.data<-format_radial(X3datuni[,4], X3datuni[,8], X3datuni[,7], X3datuni[,9], X3datuni[,1])

#Estimate causal effect of exposure 1 on Y
resX3uni<-ivw_radial(X3f.data,0.05,1,0.0001,T)

#Generate univariable radial plot for exposure 1
plotX3<-plot_radial(resX3uni,F,F,F)

png("Sim11AX3R.png",width=7,height=5.25,units="in",res=500)
plotX3
dev.off()


##############################################

###########################################
###   Radial MVMR Analyses: X1 and X2   ###
###########################################

# Select instruments for either exposure 1 or exposure 2

X1X2datuni<-sum.data[sum.data$pgammaX1hat<0.05 | sum.data$pgammaX2hat<0.05,]

#Format the X1-X2 data
X1X2f.data<-format_rmvmr(sum.data[,2:3], sum.data[,8], sum.data[,5:6], sum.data[,9], sum.data[,1])

#Estimate conditional instrument strength
strength_X1X2<- strength_rmvmr(X1X2f.data)

#Show conditional F statistic
strength_X1X2$f

#Estimate causal effects of exposure 1 and exposure 2 using Radial MVMR
X1X2res<-ivw_rmvmr(X1X2f.data,T)

#Estimate pleiotropic effects and detect outliers
pleioX1X2<-pleiotropy_rmvmr(X1X2f.data,X1X2res)

#Show Q-statistics and p-values for each SNP across all exposures
pleioX1X2$qdat

#Show global Q-statistics for all exposures
pleioX1X2$gq

X1X2plots<-plot_rmvmr(X1X2f.data,X1X2res)

png("Sim11X1X2R.png",width=7,height=5.25,units="in",res=500)
X1X2plots$p2
dev.off()

################################################
###   Radial MVMR Analyses: X1, X2, and X3   ###
################################################

# Select instruments for any exposure

X1X2X3datuni<-sum.data

#Format the X1-X2 data
X1X2X3f.data<-format_rmvmr(sum.data[,2:4], sum.data[,8], sum.data[,5:7], sum.data[,9], sum.data[,1])

#Estimate conditional instrument strength
strength_X1X2X3<- strength_rmvmr(X1X2X3f.data)

#Show conditional F statistic
strength_X1X2X3$f

#Generate conditional strength radial MVMR plot for exposure 1
strength_X1X2X3$plot[1]

#Generate conditional strength radial MVMR plot for exposure 2
strength_X1X2X3$plot[2]

#Generate conditional strength radial MVMR plot for exposure 3
strength_X1X2X3$plot[3]

#Estimate causal effects  using Radial MVMR
X1X2X3res<-ivw_rmvmr(X1X2X3f.data,T)

#Estimate pleiotropic effects and detect outliers
pleioX1X2X3<-pleiotropy_rmvmr(X1X2X3f.data,X1X2X3res)

#Show Q-statistics and p-values for each SNP across all exposures
pleioX1X2X3$qdat

#Show global Q-statistics for all exposures
pleioX1X2X3$gq

X1X2X3plots<-plot_rmvmr(X1X2X3f.data,X1X2X3res)

png("SimX1X2X3R.png",width=7,height=5.25,units="in",res=500)
X1X2X3plots$p2
dev.off()


#############################################################
###   Radial MVMR Analyses: X1, X2, and X3 with pruning   ###
#############################################################
X1X2X3fp.data<-X1X2X3f.data

outliers<-"start"

while(!is.null(outliers)){
  
  X1X2X3pres<-ivw_rmvmr(X1X2X3fp.data,F)
  pleioX1X2X3p<-pleiotropy_rmvmr(X1X2X3fp.data,X1X2X3pres)
  
  ###Prune###
  
  if(min(pleioX1X2X3p$qdat$qj_p)< 0.05){
    outliers<-pleioX1X2X3p$qdat[pleioX1X2X3p$qdat$qj_p <0.05,]$snp
    outliers<-unique(outliers)
    
    X1X2X3fp.data<-X1X2X3fp.data[!X1X2X3fp.data$SNP %in% outliers,]
    
  }else{
    outliers<-NULL
    
  }
  
}

#Estimate conditional instrument strength
strength_X1X2X3p<- strength_rmvmr(X1X2X3fp.data)

#Show conditional F statistic
strength_X1X2X3p$f

#Estimate causal effects  using Radial MVMR
X1X2X3resp<-ivw_rmvmr(X1X2X3fp.data,T)

#Estimate pleiotropic effects and detect outliers
pleioX1X2X3p<-pleiotropy_rmvmr(X1X2X3fp.data,X1X2X3resp)

#Show global Q-statistics for all exposures
pleioX1X2X3p$gq

X1X2X3plotsp<-plot_rmvmr(X1X2X3fp.data,X1X2X3resp)

png("SimX1X2X3pR.png",width=7,height=5.25,units="in",res=500)
X1X2X3plotsp$p2
dev.off()


##Plot for detecting outliers

plotdat<-pleioX1X2X3$qdat

plotdat<-plotdat[plotdat$qj_p < 0.05/(nrow(plotdat)),]

names(plotdat)[6]<- "Exposure"

plotdat$Exposure<-factor(plotdat$Exposure)

levels(plotdat$Exposure)<-c("Exposure 1","Exposure 2","Exposure 3")

F6<-ggplot(data = plotdat, aes(x = snp, y = -log10(qj_p)))+geom_point(aes(colour = Exposure))+theme_bw()+
  theme(panel.border = element_blank(),panel.grid.major = element_blank(),panel.grid.minor = element_blank(),
        axis.line = element_line(colour = "black"))+ylab(expression(-log10(p)))+xlab("Instrument")+ggtitle("Q-statistics for identified outliers")+
  geom_hline(yintercept = -log10(0.05/240))+
  scale_color_manual(values=c("Exposure 1"="#E69F00","Exposure 2"="#56B4E9","Exposure 3"="#009E73"))

png(filename = "Figure 6.png",
    width = 1920 , height = 1080, units = "px", res=250,
    bg = "white")

F6

dev.off()

#Estimate plots

#Generate unadjusted and adjusted radial MVMR plots

X1X2plots<-plot_rmvmr(X1X2f.data,X1X2res)
X1X2X3plots<-plot_rmvmr(X1X2X3f.data,X1X2X3res)
X1X2X3pplots<-plot_rmvmr(X1X2X3fp.data,X1X2X3resp)


#Generate conditional strength radial MVMR plot for exposure 1
png("X1X2_estplot.png",width=7,height=5.25,units="in",res=500)
X1X2plots$p2
dev.off()

#Generate conditional strength radial MVMR plot for exposure 1
png("X1X2X3_estplot.png",width=7,height=5.25,units="in",res=500)
X1X2X3plots$p2
dev.off()

#Generate conditional strength radial MVMR plot for exposure 1
png("X1X2X3p_estplot.png",width=7,height=5.25,units="in",res=500)
X1X2X3pplots$p2
dev.off()
```
