## Supplementary material for "Estimating and visualising multivariable Mendelian randomization analyses within a radial framework": Code,data, and additional information: application.R

```
##############################################
###   Install and load relevant packages   ###
##############################################

#install.packages("remotes")
#library(remotes)
#install_github("WSpiller/RMVMR", build_opts = c("--no-resave-data", "--no-manual"), build_vignettes = TRUE)
#install_github("WSpiller/MVMR", build_opts = c("--no-resave-data", "--no-manual"), build_vignettes = TRUE)
#install_github("WSpiller/RadialMR", build_opts = c("--no-resave-data", "--no-manual"), build_vignettes = TRUE)

#Load libraries into R
library(RMVMR)
library(RadialMR)
library(MVMR)
library(ggplot2)

#Load simulated summary data
sum.data<-read.csv("applied_data.csv",header=T)


#####################
### Univariate MR ###
#####################


#Format data for univariate MR using significant SNPs for each exposure
X1sub<-sum.data[sum.data$pgammaX1hat<5*10^-8,]
X1rad.dat<-format_radial(X1sub[,2],X1sub[,8],X1sub[,5],X1sub[,9],X1sub[,1])
X2sub<-sum.data[sum.data$pgammaX2hat<5*10^-8,]
X2rad.dat<-format_radial(X2sub[,3],X2sub[,8],X2sub[,6],X2sub[,9],X2sub[,1])
X3sub<-sum.data[sum.data$pgammaX3hat<5*10^-8,]
X3rad.dat<-format_radial(X3sub[,4],X3sub[,8],X3sub[,7],X3sub[,9],X3sub[,1])


#Perform univariate radial MR analyses for each exposure using FO weights
IVWX1A<-ivw_radial(X1rad.dat,0.05/nrow(X1rad.dat),1,0.0001,T)
IVWX2A<-ivw_radial(X2rad.dat,0.05/nrow(X2rad.dat),1,0.0001,T)
IVWX3A<-ivw_radial(X3rad.dat,0.05/nrow(X3rad.dat),1,0.0001,T)

#Format data for MVMR using X1 and X2 oriented for X1
MVMR_dat<-format_rmvmr(sum.data[,2:4],sum.data[,8],sum.data[,5:7],sum.data[,9],sum.data[,1])

#Estimate conditional instrument strength
strength_obj<- strength_rmvmr(MVMR_dat)

#Show conditional F statistics
strength_obj$f

#Generate conditional strength radial MVMR plot for HDL
strength_obj$plot[1]

#Generate conditional strength radial MVMR plot for LDL
strength_obj$plot[2]

#Generate conditional strength radial MVMR plot for TG
strength_obj$plot[3]

#Format data for MVMR using X1 and X2 oriented for X1
MVMR_dat<-format_rmvmr(sum.data[,2:4],sum.data[,8],sum.data[,5:7],sum.data[,9],sum.data[,1])

#Estimate conditional instrument strength
strength_obj<- strength_rmvmr(MVMR_dat)

#Show conditional F statistics
strength_obj$f

#Generate conditional strength radial MVMR plot for LDL
strength_obj$plot[1]

#Generate conditional strength radial MVMR plot for HDL
strength_obj$plot[2]

#Generate conditional strength radial MVMR plot for TG
strength_obj$plot[3]

#Estimate causal effects  using Radial MVMR
mvmrres<-ivw_rmvmr(MVMR_dat,T)

#Estimate pleiotropic effects and detect outliers
pleiomvmr<-pleiotropy_rmvmr(MVMR_dat,mvmrres)

#Show global Q-statistics for all exposures
pleiomvmr$gq

#Generate unadjusted and adjusted radial MVMR plots
mvmrplots<-plot_rmvmr(MVMR_dat,mvmrres)
mvmrplots$p1
mvmrplots$p2

#############################################################
###   Radial MVMR Analyses: X1, X2, and X3 with pruning   ###
#############################################################


mvmrp.data<-MVMR_dat

outliers<-"start"

while(!is.null(outliers)){
  mvmrpres<-ivw_rmvmr(mvmrp.data,F)
  pleiomvmrp<-pleiotropy_rmvmr(mvmrp.data,mvmrpres)
  ###Prune###
  if(min(pleiomvmrp$qdat$qj_p)< 0.05){
    outliers<-pleiomvmrp$qdat[pleiomvmrp$qdat$qj_p <0.05,]$snp
    outliers<-unique(outliers)
    mvmrp.data<-mvmrp.data[!mvmrp.data$SNP %in% outliers,]
  }else{
    outliers<-NULL
  }
}

#Estimate conditional instrument strength
strength_objp<- strength_rmvmr(mvmrp.data)
strength_objp$f

#Generate conditional strength radial MVMR plot for exposure 1
strength_objp$plot[1]

#Generate conditional strength radial MVMR plot for exposure 2
strength_objp$plot[2]

#Generate conditional strength radial MVMR plot for exposure 3
strength_objp$plot[3]

#Estimate causal effects  using Radial MVMR
mvmrpres<-ivw_rmvmr(mvmrp.data,T)

#Show global Q-statistics for all exposures
pleiomvmrp$gq

#Generate unadjusted and adjusted radial MVMR plots
mvmrpplots<-plot_rmvmr(mvmrp.data,mvmrpres)

#Outliers
MVMR_dat[!MVMR_dat$SNP %in% mvmrp.data$SNP,]$SNP

png(filename = "Figure 9D.png",
    width = 1400 , height = 1200, units = "px", res=300,
    bg = "white")

mvmrplots$p2

dev.off()
```
