## Supplementary material for "Estimating and visualising multivariable Mendelian randomization analyses within a radial framework": Code,data, and additional information: RMVMR_supplementary.docx

**Supplementary Material (updated 09/01/21)**

### A1: Derivation of the ratio estimate

For $i\in\left( 1,2,\ldots,N \right)$ observations including $j\in(1,2,\ldots,J)$ SNPs, a set of SNPs $l\in\left( 1,2,\ldots L \right)$ where $j\notin l$, $k\in\left( 1,2,\ldots,K \right)$ exposures, and a set of exposures $m\in\left( 1,2,\ldots,m \right)$ for which $k\notin m$:

Data Generating Model

$G_{ji}\sim\theta_{j0}+\sum_{l=1}^{L} \theta_{jl}G_{l}+\epsilon_{Gji}$	(A2.1)

$U_{i}=\pi_{0}+\sum_{j=1}^{J} \pi_{j}G_{j}+\epsilon_{U_{i}}$	(A2.2)

$X_{ki}=\gamma_{k0}+\sum_{j=1}^{J} \gamma_{kj}G_{j}+\gamma_{k\left( J+1 \right)}U_{i}+\sum_{m=1}^{M} \delta_{km}X_{m}+\epsilon_{Xki}$	(A2.3)

$Y_{i}=\beta_{0}+\sum_{k=1}^{K} \beta_{k}X_{k}+\sum_{j=1}^{J} \alpha_{j}G_{j}+\beta_{\left( K+1 \right)}U_{i}+\epsilon_{Yi}$	(A2.4)

Linear regression of $Y$ on all variables

$Y_{i}=\hat{\beta}_{0}+\sum_{k=1}^{K} \hat{\beta}_{k}X_{k}+\sum_{j=1}^{J} \hat{\alpha}_{j}G_{j}+\hat{\beta}_{\left( K+1 \right)}U_{i}+\hat{\epsilon}_{Yi}$	(A2.5)

Simple linear regression of $G_{j}$ on $G_{l}$

$G_{ji}=\tilde{\theta}_{lj0}+{\tilde{\theta}_{jl}G}_{li}+\tilde{\epsilon}_{Gjli}$	(A2.6)

Simple linear regression of $U$ on $G_{j}$

$U_{i}=\tilde{\pi}_{j0}+\tilde{\pi}_{j}G_{j}+\tilde{\epsilon}_{Uji}$	(A2.7)

Linear regression of $Y$ on all variables

$X_{ki}=\hat{\gamma}_{k0}+\sum_{j=1}^{J} \hat{\gamma}_{kj}G_{j}+\hat{\gamma}_{k\left( J+1 \right)}U_{i}+\epsilon_{Xki}$	(A2.8)

Simple linear regression of $X_{k}$ on $G_{j}$

$X_{k}=\tilde{\gamma}_{kj0}+\tilde{\gamma}_{kj}G_{j}+\tilde{\epsilon}_{Xkji}$	(A2.9)

Total effect of $G_{j}$ on exposure $X_{k}$

$\tilde{\gamma}_{kj}=\hat{\gamma}_{kj}+\sum_{l=1}^{L} \hat{\gamma}_{kl}\hat{\theta}_{jl}+\sum_{m=1}^{M} \delta_{km}\tilde{\gamma}_{kj}+\gamma_{k\left( J+1 \right)}\tilde{\pi}_{j}$	(A2.9)

Reduced form simple linear regression of Y on instrument $G_{j}$

$Y_{i}=\tilde{\Gamma}_{j0}+\tilde{\Gamma}_{ji}+\tilde{\eta}_{i}$	(A2.10)

Total effect of $G_{j}$ on $Y$

$\tilde{\Gamma}_{j}=\sum_{k=1}^{K} \hat{\beta}_{k}\tilde{\gamma}_{kj}+\hat{\alpha}_{j}+\sum_{l=1}^{L} \hat{\alpha}_{l}\tilde{\theta}_{lj}+\hat{\beta}_{K+1}\tilde{\pi}_{j}$	(A2.11)

Wald ratio estimate $\hat{\beta}_{kj}$ for instrument $G_{j}$

$\hat{\beta}_{kj}=\frac{\tilde{\Gamma}_{j}}{\tilde{\gamma}_{kj}}=\frac{\sum_{k=1}^{K} \hat{\beta}_{k}\tilde{\gamma}_{kj}+\hat{\alpha}_{j}+\sum_{l=1}^{L} \hat{\alpha}_{l}\tilde{\theta}_{lj}+\hat{\beta}_{K+1}\tilde{\pi}_{j}}{\hat{\gamma}_{kj}+\sum_{l=1}^{L} \hat{\gamma}_{kl}\hat{\theta}_{jl}+\sum_{m=1}^{M} \delta_{km}\tilde{\gamma}_{mj}+\gamma_{k\left( J+1 \right)}\tilde{\pi}_{j}}$	(A2.12)

$\frac{\tilde{\Gamma}_{j}}{\tilde{\gamma}_{kj}}=\hat{\beta}_{k}+\frac{\sum_{m=1}^{M} \hat{\beta}_{m}\tilde{\gamma}_{mj}+\hat{\alpha}_{j}+\sum_{l=1}^{L} \hat{\alpha}_{l}\tilde{\theta}_{lj}+\hat{\beta}_{K+1}\tilde{\pi}_{j}}{\hat{\gamma}_{kj}+\sum_{l=1}^{L} \hat{\gamma}_{kl}\hat{\theta}_{jl}+\sum_{m=1}^{M} \delta_{km}\tilde{\gamma}_{mj}+\gamma_{k\left( J+1 \right)}\tilde{\pi}_{j}}$	(A2.13)

MVMR1: $\tilde{\gamma}_{kj}=\hat{\gamma}_{kj}+\sum_{l=1}^{L} \hat{\gamma}_{kl}\hat{\theta}_{jl}+\sum_{m=1}^{M} \delta_{km}\tilde{\gamma}_{mj}\neq0$

MVMR2: $\pi_{j}=0$

MVMR3: $\alpha_{j}=0$

$\frac{\tilde{\Gamma}_{j}}{\tilde{\gamma}_{kj}}=\hat{\beta}_{k}+bias\left( \frac{\sum_{m=1}^{M} \hat{\beta}_{m}\tilde{\gamma}_{mj}}{\hat{\gamma}_{kj}+\sum_{l=1}^{L} \hat{\gamma}_{kl}\hat{\theta}_{jl}+\sum_{m=1}^{M} \delta_{km}\tilde{\gamma}_{mj}} \right)$	(A2.14)

$\frac{\tilde{\Gamma}_{j}}{\tilde{\gamma}_{kj}}=\hat{\beta}_{k}+bias\left( \frac{\sum_{m=1}^{M} \hat{\beta}_{m}\tilde{\gamma}_{mj}}{\tilde{\gamma}_{kj}} \right)$	(A2.15)

When the MVMR assumptions are satisfied, the univariable ratio bias term using a single instrument with respect to exposure $X_{k}$ is equal to the sum of the additional effects of exposures $X_{m}$ divided by the total effect of the SNP on exposure $X_{k}$. This is adjusted for when including information on exposures $X_{m}$ within an MVMR model.

### A2: Simulation study: Further information

Univariable Radial MR Plots

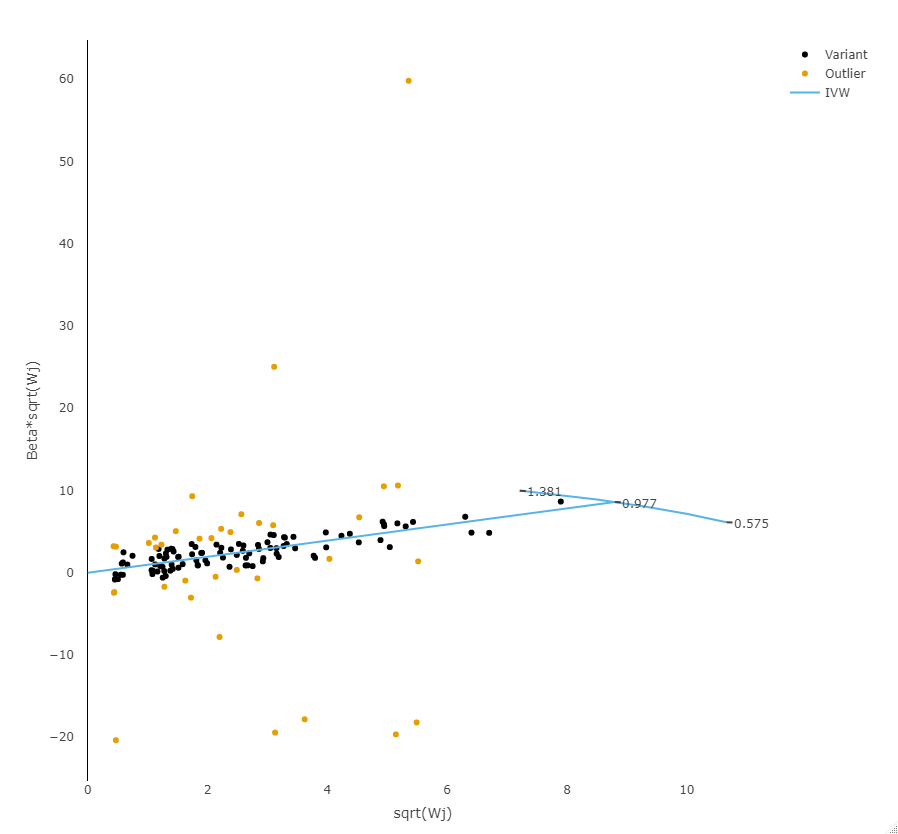

***Figure A2.1***: A Radial MR plot showing the estimated causal effect of exposure $X_{1}$ upon outcome $Y$. Observations represent the ratio estimate for each SNP robustly associated with exposure $X_{1}$.

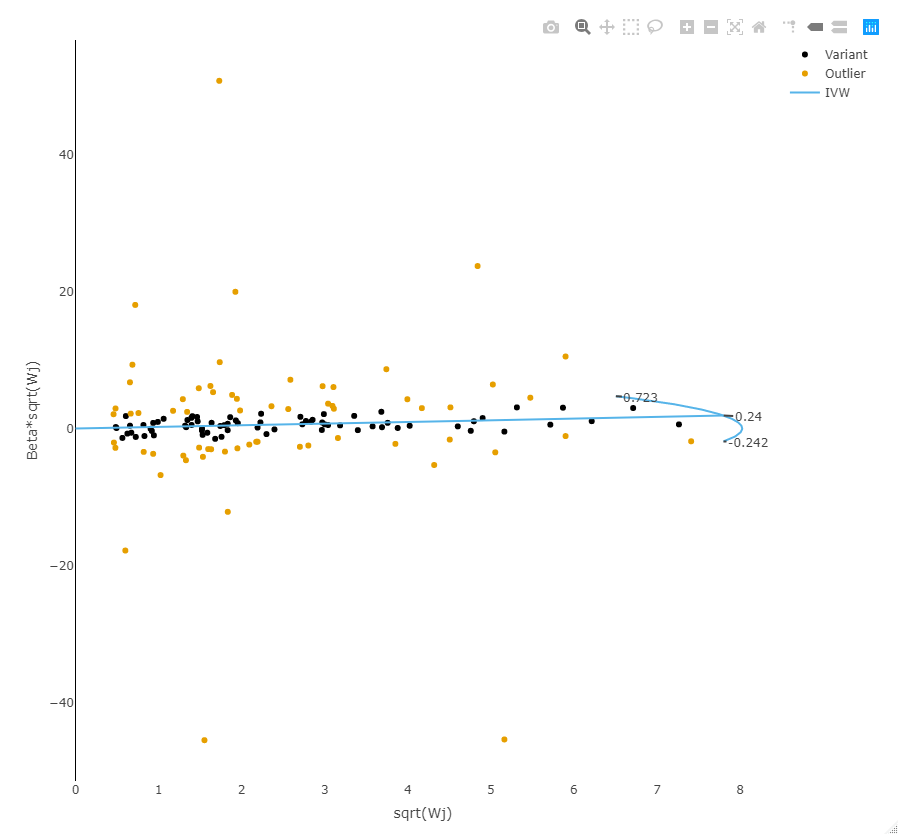

***Figure A2.2***: A Radial MR plot showing the estimated causal effect of exposure $X_{2}$ upon outcome $Y$. Observations represent the ratio estimate for each SNP robustly associated with exposure $X_{2}$.

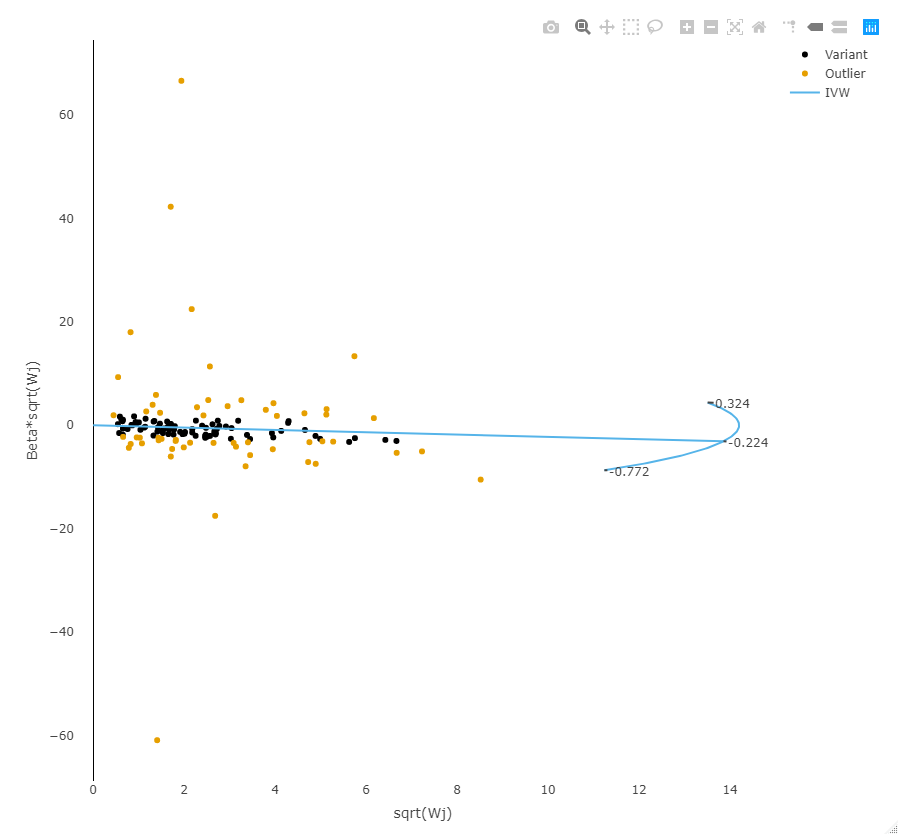

***Figure A2.3***: A Radial MR plot showing the estimated causal effect of exposure $X_{3}$ upon outcome $Y$. Observations represent the ratio estimate for each SNP robustly associated with exposure $X_{3}$.

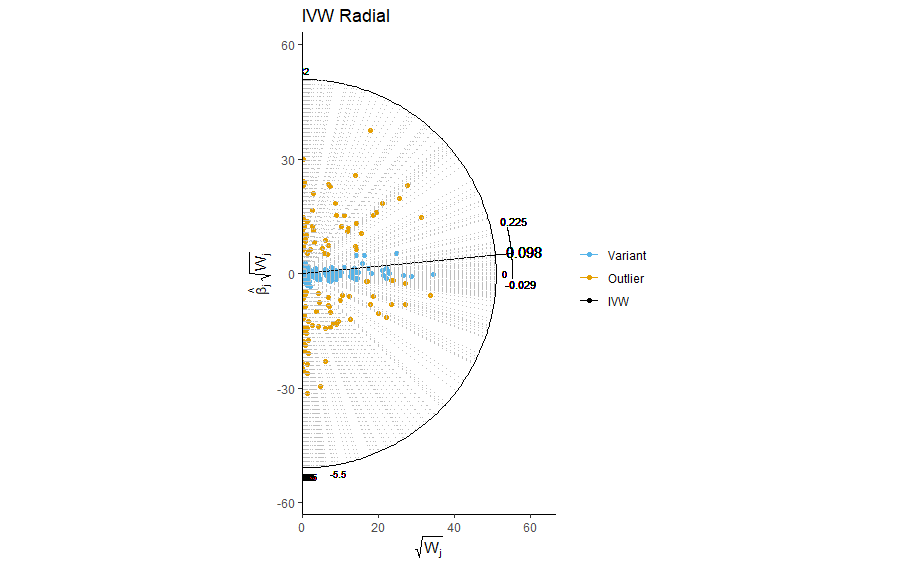

***Figure A2.4***: A Radial MR plot showing heterogeneity indicative of conditional instrument strength for exposure $X_{1}$

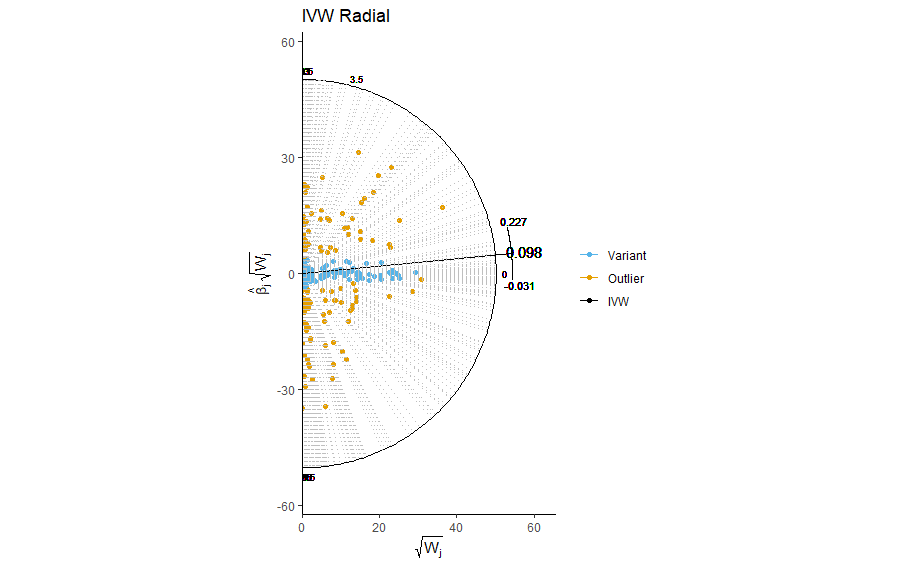

***Figure A2.5***: A Radial MR plot showing heterogeneity indicative of conditional instrument strength for exposure $X_{2}$

### A3: Further information on applied analyses

Univariate Radial MR Plots

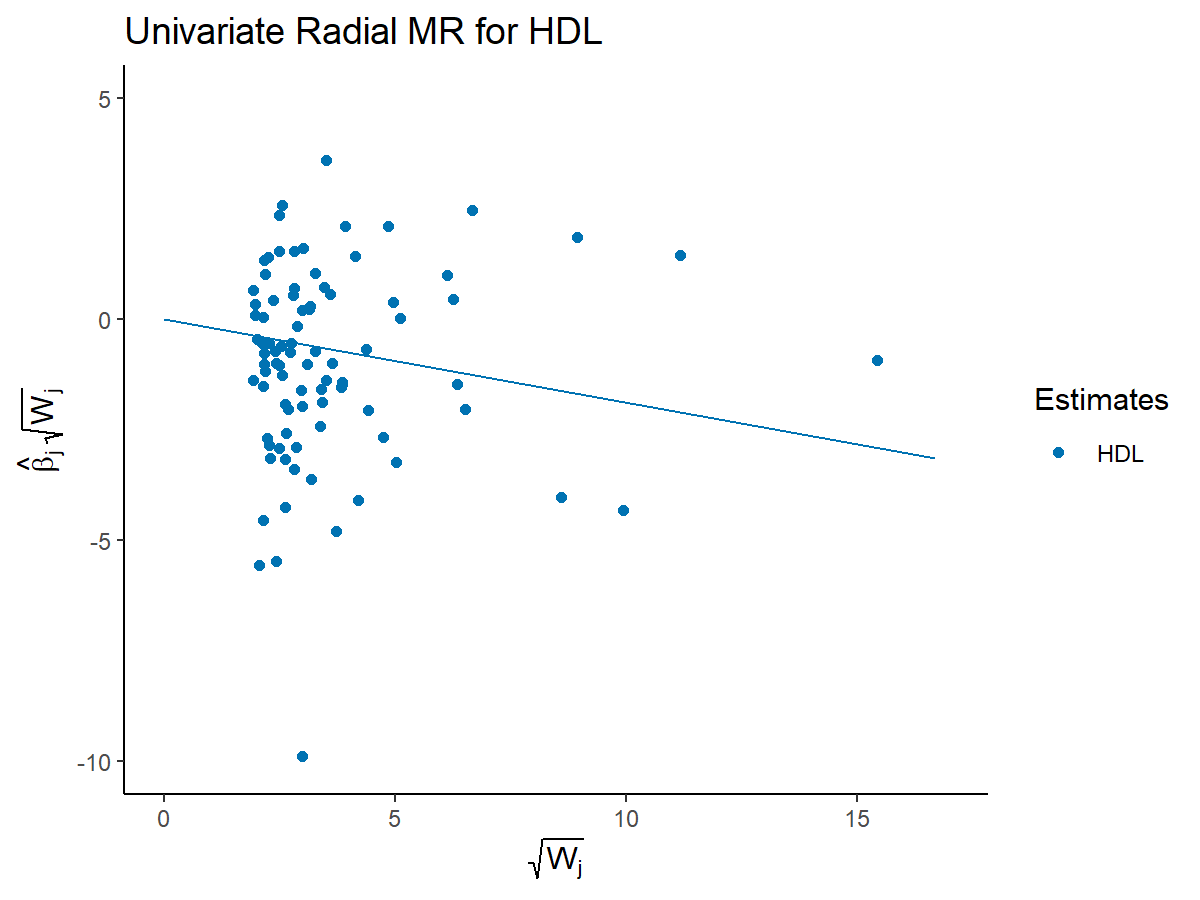

***Figure A3.1***: A Radial MR plot showing the estimated causal effect of HDL upon CHD. Observations represent the ratio estimate for each SNP robustly associated with HDL.

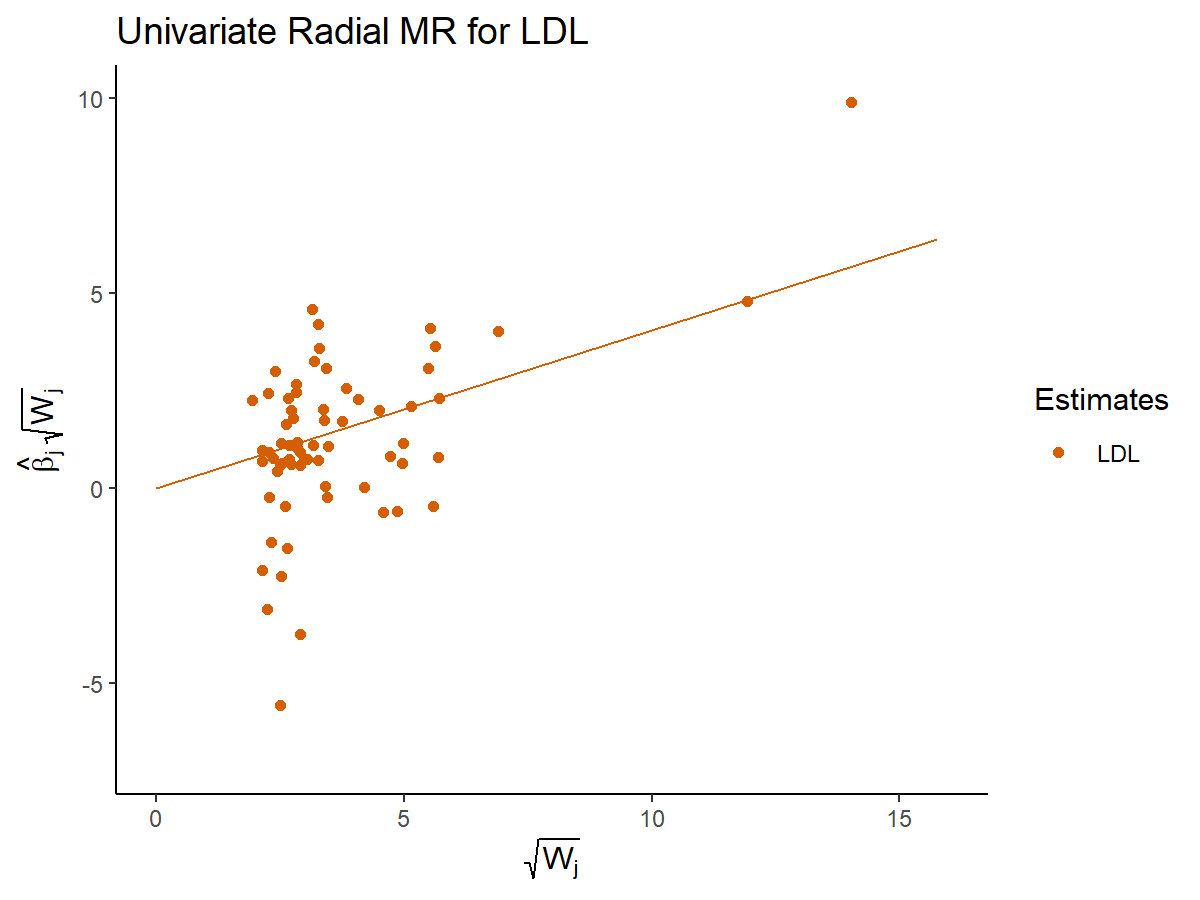

***Figure A3.2***: A Radial MR plot showing the estimated causal effect of LDL upon CHD. Observations represent the ratio estimate for each SNP robustly associated with LDL.

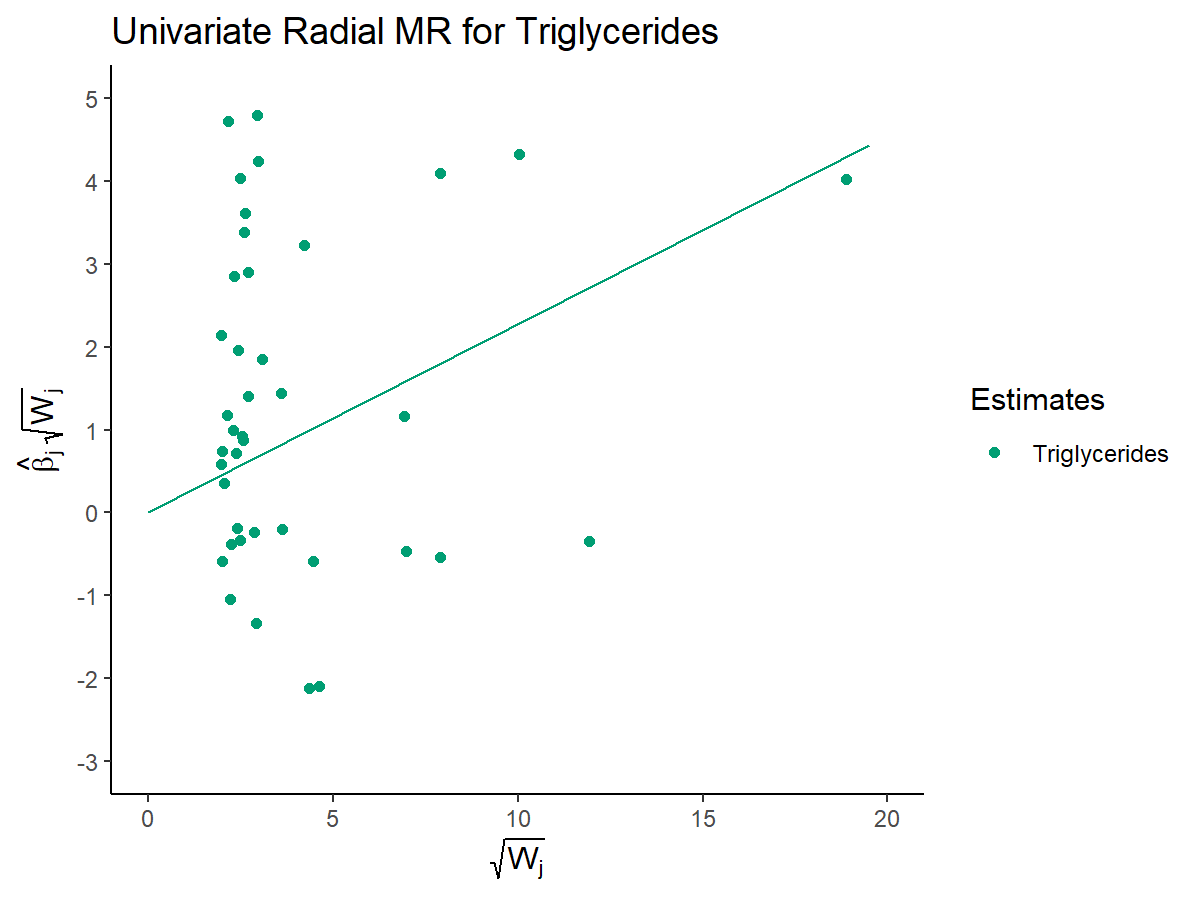

***Figure A3.3***: A Radial MR plot showing the estimated causal effect of triglycerides upon CHD. Observations represent the ratio estimate for each SNP robustly associated with triglycerides.

SNPs identified as outliers in radial MVMR analysis

rs11065987

rs11153594

rs11244084

rs1250229

rs1260326

rs12740374

rs12801636

rs205262

rs2068888

rs2288912

rs2390536

rs267733

rs3106166

rs35633988

rs4530754

rs6567160

rs998584
